# Evaluation of the genome-informed risk assessment (GIRA) approach from eMERGE in an independent health system

**DOI:** 10.64898/2026.02.25.26347123

**Authors:** Sandra Lapinska, Xinzhe Li, Ravi Mandla, Zhuozheng Shi, Veronica Tozzo, Alexander Flynn-Carroll, Marylyn D. Ritchie, Daniel J. Rader, Penn Medicine Biobank, Bogdan Pasaniuc

## Abstract

The Genome Informed Risk Assessment (GIRA) from eMERGE is an ongoing pragmatic prospective study designed to implement and evaluate genomic precision medicine across diverse clinical settings. Here, we examine the utility of the high-risk criteria from GIRA in assessing health risk through a retrospective evaluation of 9 adult conditions in a health system independent of eMERGE using the Penn Medicine Biobank (PMBB, n=48,279). We find a large proportion of patients - 30.4% (n=14,676) - labeled as high-risk based on the genomic components (monogenic and polygenic) of GIRA. Stratifying by ancestry revealed significant differences in high-risk classification, with higher rates in African/African American (56.6% vs. 50.1%, p=7.43×10^-36^) and lower rates in East (42.0%) and South Asian (40.0%) ancestries when considering all three high-risk components of GIRA. Enrichments of high-risk individuals were observed in the highest quartile of social deprivation index highlighting contextual influences on high-risk measures. The polygenic component of GIRA was a good predictor of prevalent cases. GIRA predicted incident disease albeit with lower accuracies for some conditions. We find demographic compositions of high-risk patients differed from the incident cases for certain conditions; for example, high-risk AFIB individuals where enriched for European ancestries in contrast with incident AFIB cases that were enriched for AFR ancestries. Overall, our results show the accuracy of GIRA’s high-risk criteria as a biomarker to stratify high-risk patients for precision medicine and highlight its potential impact on the health system if implemented at scale.

## Introduction

Current screening practices for identifying individuals at high-risk of disease rely on clinical risk factors and family history (FH), with assessment of monogenic variants being incorporated for select cancer and cardiometabolic conditions as it can confer several-fold increased disease risk for carriers^1–5^. Family history may reflect rare pathogenic variants of heredity, shared exposures to environmental and lifestyle factors, as well as common genetic susceptibility, thus providing an indirect measure of overall genetic predisposition to disease^6–10^. More recently, polygenic risk scores (PGS) that aggregate the effects of many common variants into a single measure of risk have demonstrated great potential in risk stratification as individuals in the upper tail of distribution can attain risk equivalent to clinical risk factors and monogenic variants^4,11–13^. Thus, combining PGS with family history and monogenic risk provides a more comprehensive genetic risk assessment for common, complex diseases, enabling earlier identification of high-risk individuals who may benefit from preventative interventions^1^. Additionally, individuals carrying the same monogenic variant demonstrate wide variation in disease expression and penetrance^14^. Considering polygenic background improves risk estimation, as it can modify the penetrance and expressivity of monogenic variants^13^. Moreover, for most conditions, a positive family history with a high PGS increases disease risk, whereas a low PGS can offset the risk associated with a positive family history^6,15^. Recognizing that each of these genetic risk factors provides independent and complementary information into inherited disease susceptibility, the Electronic Medical Records and Genomics (eMERGE) Network^16,17^ has developed the genome-informed risk assessment (GIRA) report as part of their ongoing prospective study across multiple clinical sites. eMERGE deployed cross-ancestry and ancestry-calibrated PGS, together with monogenic risk and family history, across 11 conditions (9 adult) to evaluate health outcomes among high-risk individuals and inform best practices for large-scale genomic medicine^1,18,19^. High PGS status for each condition was selected to correspond to an expert specified, clinically meaningful level of risk, established by eMERGE, such as an odds ratio ≥ 2 or positive family history^20^.

Retrospective evaluation of the high-risk criteria from GIRA in healthcare systems independent of eMERGE, where the patient populations may differ from those used to calibrate GIRA, is a critical step towards broad adoption of genomic precision medicine^21^. Specifically, GIRA’s genomic and family history components of risk prediction may show diminished performance in systems outside eMERGE due to differences in ancestry proportions, environmental exposures, disease prevalence, or case ascertainment^1,18,22^. For instance, in the University of Pennsylvania Health System (UPHS), 55.0% of the patient population self-reported as ‘White’, 18.2% as ‘African American’, and 4.1% as ‘Asian’, with 4.7% self-reporting ethnicity as ‘Hispanic’^23^. The Penn Medicine Biobank (PMBB), an electronic health record (EHR)-linked biobank embedded within the UPHS, includes 23.2% individuals of African/African American (AFR) ancestry, the largest representation within any single-institutional medical biobank in the US, providing a unique opportunity to validate the population-specific performance of each risk component of GIRA. More specifically, even though PMBB has a higher proportion of individuals who self-reported as ‘White’ (66.8%) and ‘African American’ (24.6%), it is largely representative of the UPHS and a valuable patient population for validation. The clinical utility of the GIRA report also depends on its ability to identify individuals at high-risk of developing the condition in the future as it enables more targeted interventions^24–26^. Evaluation of GIRA for prevalent conditions may overstate GIRA’s predictive ability, since a strong association with prevalent conditions does not necessarily reflect how well the tool predicts future condition events^27^. For instance, PGS strongly associate with prevalent conditions but have lower predictive accuracy in incident conditions^28^. Cox proportional hazard models for incidence disease (defined as individuals that develop condition after being evaluated for GIRA high-risk outcomes), on the other hand, directly quantify prospective risk by estimating how risk components within genetic risk assessments influence the likelihood of developing the disease, providing a more clinically meaningful assessment of prediction. To date, however, no studies have longitudinally evaluated each of the GIRA high-risk components in a system independent of eMERGE for incident disease prediction.

We leveraged longitudinal data in PMBB to investigate the predictive accuracy of each GIRA components (monogenic, polygenic, family history) to assess their ability in accurately capturing high-risk patients^23^. Prevalent cases were defined as any diagnosis recorded before, at, or after entry in biobank (baseline), whereas incident cases were diagnosed after PMBB entry only. We implemented validated phenotype algorithms for nine adult-onset conditions - atrial fibrillation (AFIB), breast cancer (BC), chronic kidney disease (CKD), coronary heart disease (CHD), hypercholesterolemia (HC), obesity, prostate cancer (PC), type 2 diabetes (T2D), and colorectal cancer (CC) – and evaluated the ability of GIRA to identify disease cases. In total, 50.1.% (17,747) of unique PMBB patients were labeled as high-risk due to polygenic, monogenic, and/or family history risk for at least one condition (28.7%, 2.3%, and 28.8%, respectively). Of these, 30.4% (n=14,676) are due to the genomic component of GIRA (monogenic and polygenic). Stratifying PMBB patients by ancestry found significant differences in high-risk proportions when compared to all patients, most notably AFR individuals having a high-risk GIRA due to high PGS and family history at elevated rates (e.g. high-risk PGS: 34.2% vs 28.7%, p=9.78×10^-30^) than in other ancestries suggesting a need for ancestry-specific calibration. Lower GIRA-risk rates in East Asian ancestry were driven solely by family history suggesting a larger impact from factors like reporting pattern variability than differences in estimated genetic risk. Of the 17,747 high-risk GIRA individuals, 65.0% were diagnosed with at least one condition before PMBB recruitment and 28.3% developed at the condition after PMBB entry (defined as incident cases). Effect sizes of high PGS to identify prevalent cases were largely consistent with those reported by eMERGE ranging from 2.28 to 3.97 for BC and T2D. For prediction of incident disease, hazard ratios (HRs) showed significant associations across all GIRA risk components with hazard ratios varying from 1.31 (95% CI: 1.05-1.63) for AFIB to 2- to 5-fold for all other conditions. High-risk GIRA individuals were of similar ancestry composition to incident cases across all conditions except AFIB, CKD, and obesity suggesting potential for miscalibration across ancestries. Finally, elevated GIRA-risk rates were observed in the highest quartile of social deprivation index for high PGS and family history, whereas lower rates were observed for monogenic risk highlighting the potential miscalibration of GIRA due to environmental factors or healthcare access. In summary, GIRA risk components showed considerable promise for identifying high-risk individuals in an independent evaluation informed by eMERGE. However, its underperformance for certain conditions highlights the need for condition- and context-specific validation and refinement to ensure equitable clinical implementation in real-world healthcare systems.

## Results

### Phenotyping nine adult GIRA conditions in the Penn Medicine Biobank (PMBB)

We analyzed 48,279 PMBB patients aged 18 to 75 years at sample collection. Majority of these individuals were genetically similar to European American (EUR) (71.7%) and African/African American (AFR) (23.2%) populations (**Figure 1; Table S1; Methods**). PMBB has one of the largest sample size of AFR individuals among single-institutional medical biobanks, making it valuable for evaluating the generalizability of GIRA across diverse populations^29–31^. To closely align our analysis with eMERGE, we established case and control status for nine adult GIRA conditions using validated phenotyping algorithms from the Phenotype Knowledgebase (PheKB) when applicable; otherwise, the Chronic Condition Warehouse or the Polygenic Risk Methods Development (PRIMED) Consortium were used (**Methods**). We observed higher prevalence estimates than previously reported population-based estimates for all traits except BC, CC, PC, and obesity (**Figure 1; Table S1, S3**). For example, population-based prevalence estimates for CHD, T2D, and AFIB are approximately 6.4%^32^, 6.28%^33^, and 13.7%^34^, respectively, significantly lower than compared to 13.7%, 14.5%, and 17.2% in PMBB. In contrast, CKD prevalence in PMBB (11.5%) was lower than reported population estimates of 14%^35^. Comparison of disease prevalence between PMBB and the original GIRA validation cohort revealed statistically significant higher prevalence in PMBB for 3 conditions and lower prevalence for 5 conditions (**Table S3**). For example, prevalence was significantly lower in PMBB than in eMERGE for AFIB (17.2% vs. 23.0%; P = 1.13×10^-1O4^) and CKD ( 11.5% vs. 17.5%; P = 6.95×10^-22O^) whereas CHD (13.7% vs. 9.2%; P = 1.41×10^-146^) and HC (12.1% vs. 8.1%; P = 4.05×10^-48^) were more prevalent in PMBB. Prevalence estimates in PMBB generally fell between population-based estimates and those observed in eMERGE. Higher prevalence for most conditions is likely due to volunteer bias in biobank participants which has been shown to distort prevalence estimates relative to the target population^36,37^. As expected, incidence estimates, defined as receiving diagnosis after sample collection in PMBB, were significantly lower than corresponding prevalence estimates. Observation time was defined as the earliest of the following times: initial disease diagnosis, last clinical encounter, or death. Incidence estimates ranged from 0.5% for CC to 7.2% for obesity. PMBB incidence estimates differed to population-level incidence; however, AFIB has reported incidence rates of 23.7 per 1000 person-years (PY) over 1-year follow-up^34^ (compared with 5.69 per 1000PY over 1-year follow-up in PMBB) while BC has 130.8 per 100,000 women^38^ (compared with 227.0 per 100,000 women per year in PMBB), reflecting cohort-specific differences in ascertainment (**Figure 1; Table S2, S3**).

**Figure 1.**
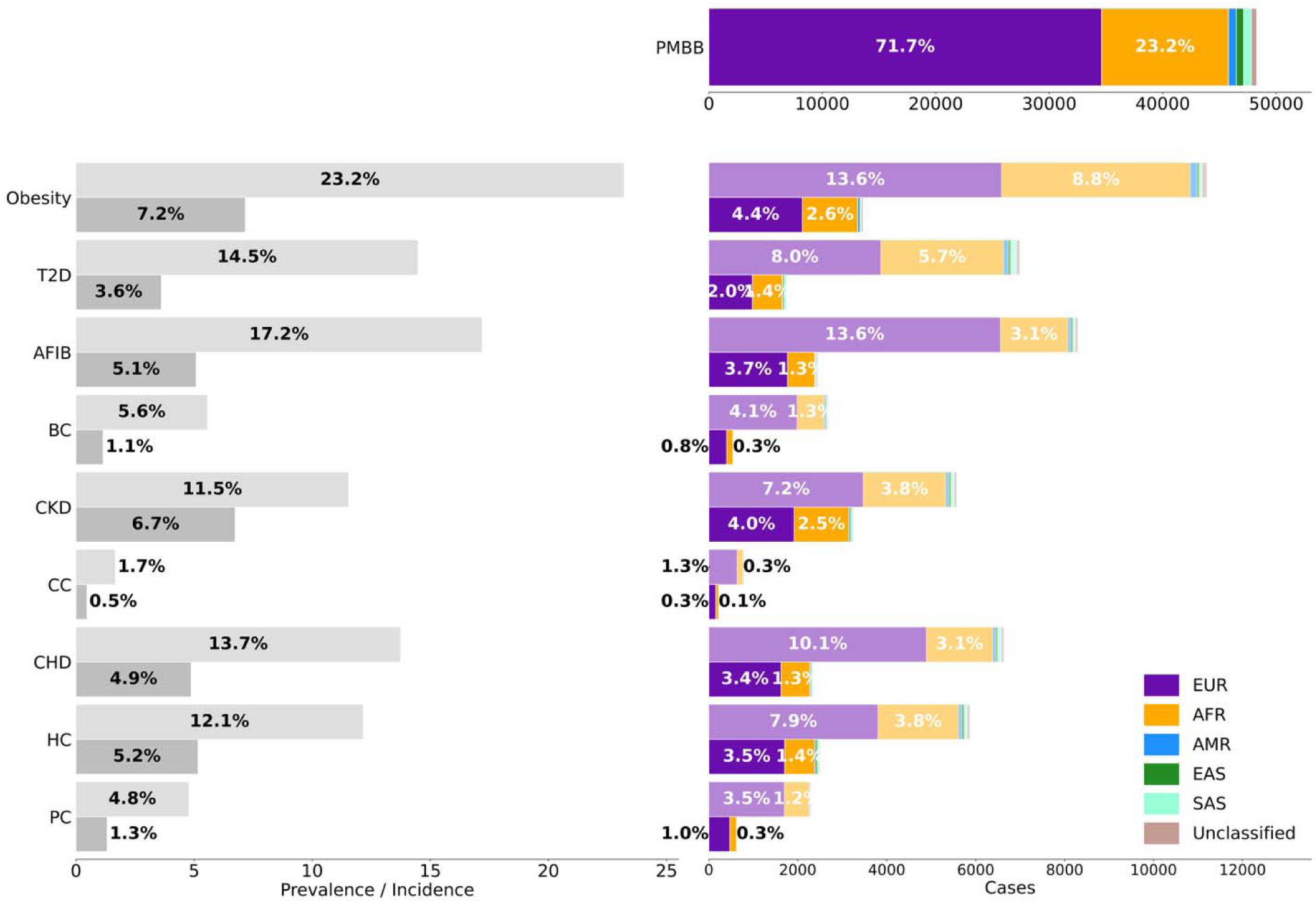
Distribution of nine adult GIRA conditions across ancestry, age, and sex in PMBB. Left panel shows the prevalence (defined as participants diagnosed at dates prior to, at, and post enrollment) compared to the incidence (defined as participants receiving diagnosis after sample collection) of each condition within PMBB. Right shows the distribution of cases by genetically defined ancestry groupings with distribution in the overall biobank at the top. Bars are annotated with ancestry proportions for participants of EUR or AFR ancestry only, as these groups contain the majority of the participants. The ancestry proportions are shown relative to the total PMBB participants included in the analysis. We included all PMBB participants aged 18 to 75 at sample collection who were not enrolled in the Penn Medicine Cancer Risk Evaluation Program. Abbreviations: GIRA: Genome Informed Risk Assessment, PMBB: Penn Medicine Biobank, T2D: Type 2 Diabetes, AFIB: Atrial Fibrillation, BC: Breast Cancer, CKD: Chronic Kidney Disease, CC: Colorectal Cancer, CHD: Coronary Heart Disease, HC: Hypercholesterolemia, PC: Prostate Cancer, EUR: European American ancestry, AFR: African/African American ancestry, AMR: Admixed American ancestry, EAS: East Asian ancestry, SAS: South Asian ancestry

### Demographic characteristics of prevalent and incident disease in PMBB

Next, to gain insights into ascertainment effects in biobank enrollment, we compared ancestry, age and sex enrichments in prevalent versus incident cases for each condition. Overall, we observed similar ancestry distributions in prevalent vs incident cases across conditions with few differences. We found AFR individuals were enriched in both prevalent and incident obesity, T2D, CKD, CHD, and PC whereas EUR individuals were enriched in BC and CC, largely in-line with existing epidemiological studies^39–43^. Differences in ancestry enrichment patterns were observed between prevalent and incident cases for AFIB and HC. We found an enrichment of AFR individuals in incident AFIB (OR: 1.14 (95% CI: 1.03-1.26)) whereas prevalent AFIB was enriched in EUR individuals (OR: 1.39 (95% CI: 1.31-1.48)). In contrast, incident HC was enriched for EUR individuals (OR: 1.10 (95% CI: 1.00-1.21)) while prevalent HC was enriched for AFR individuals (OR: 1.16 (95% CI: 1.08-1.24)). These enrichment patterns suggest possible participation and recruitment biases in PMBB that could be impacted by factors like survival bias, disease severity, healthcare access and utilization, or issues regarding inclusivity (**Figure 2; Table S7**)^44^. Biobanks are known to suffer from inclusion bias with several studies comparing participants to the background population using publicly available census data^45,46^. Next, we found incident T2D and BC were enriched for older individuals, specifically those aged 45-65 and aged 65-100 (e.g. incident T2D OR: 1.27 (95% CI: 1.15-1.41) vs prevalent T2D OR: 0.68 (95% CI: 0.63-0.72)), compared to prevalent cases. Incident CC also showed enrichment for individuals aged 65-100. Prevalent AFIB, CKD, CC, CHD, and PC were enriched for individuals aged 45-65 whereas incident cases were not (**Supplementary Figure 1; Table S8**). These findings highlight that incident disease tends to skew toward older individuals, likely reflecting longer survival trends among adults aged 60 or older with common, not immediately fatal conditions like T2D, AFIB, or HC^47,48^. Conversely, enrichment among individuals aged 18-45 was observed only for prevalent CC (OR: 1.25 (95% CI: 0.99-1.58)), mirroring trends of increasing diagnoses among individuals under age 55^49^, likely driven by earlier screening practices and resulting in greater biobank enrollment among younger individuals with elevated CC risk factors. Lastly, all conditions were enriched for males except prevalent and incident obesity and HC, consistent with previous epidemiological studies^50–54^, aside from CKD, which is more prevalent globally among women^35,55^. This may be due to potential under-screening and underdiagnosis of CKD among women within healthcare systems^56^. BC and PC were not evaluated for sex enrichment as GIRA returns results to self-reported sex at birth of female and male, respectively (**Supplementary Figure 2; Table S9**). Together, these findings demonstrate how ancestry, age, and sex enrichments reflect known epidemiologic patterns while also revealing potential ascertainment biases between prevalent and incident cases that need to be considered in downstream analyses.

**Figure 2.**
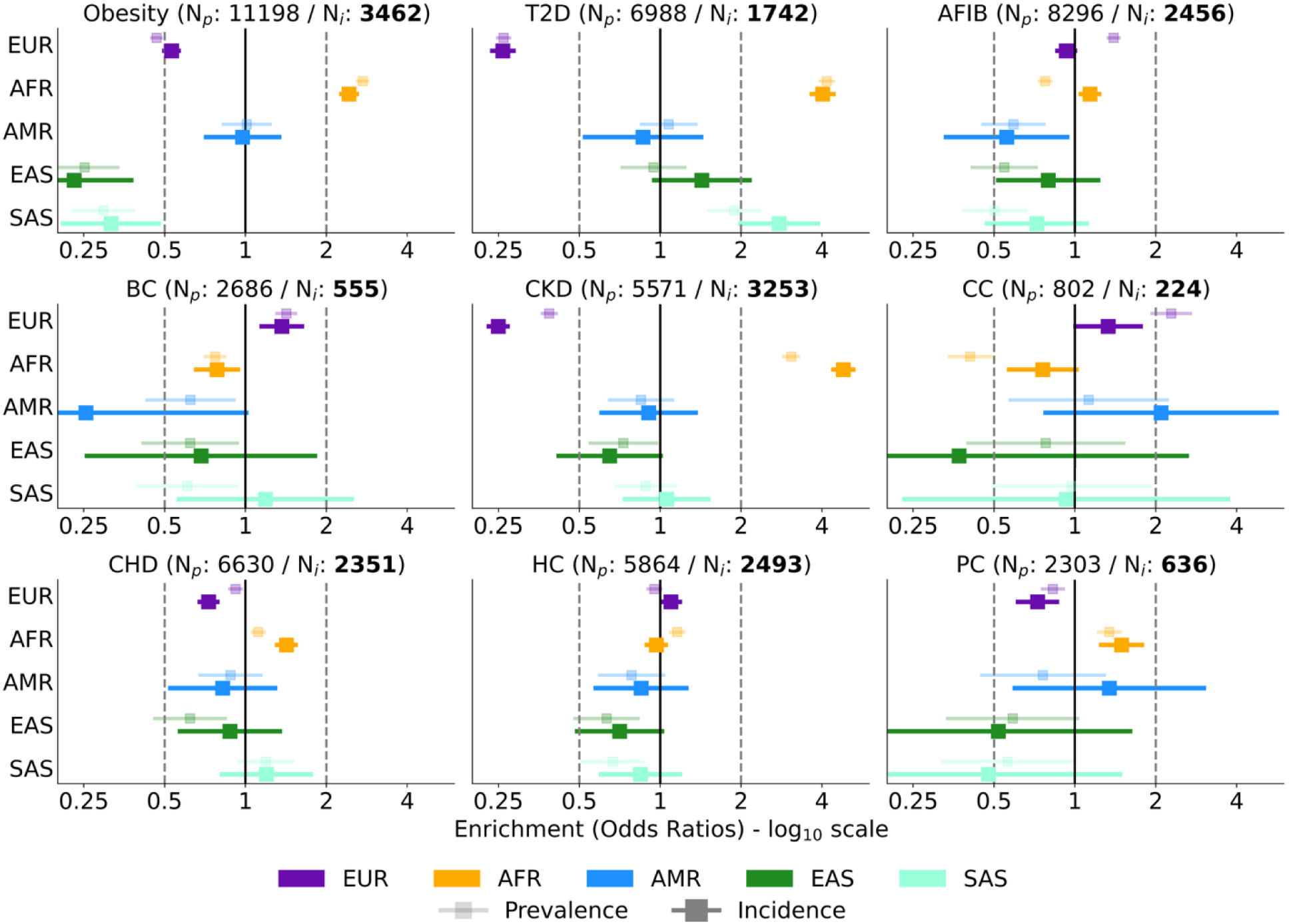
Ancestry enrichments within nine conditions as compared to PMBB participants. Enrichment within prevalent (lighter shade) versus incident (darker shade) cases across the nine adult GIRA conditions is estimated using odds ratios and their standard errors. N_p_ (N_i_) denotes the number of prevalent (incident) cases for each condition among biobank participants aged 18 to 75 at the time of sample collection who were not enrolled in the Penn Medicine Cancer Risk Evaluation Program (N_total_=48,279). Odds ratios are plotted on the log_10_ scale. Cutoff bars extend beyond the displayed range and were truncated at x-axis limits of 0.25 and 6 for visualization. Abbreviations: GIRA: Genome Informed Risk Assessment, PMBB: Penn Medicine Biobank, T2D: Type 2 Diabetes, AFIB: Atrial Fibrillation, BC: Breast Cancer, CKD: Chronic Kidney Disease, CC: Colorectal Cancer, CHD: Coronary Heart Disease, HC: Hypercholesterolemia, PC: Prostate Cancer, EUR: European American ancestry, AFR: African/African American ancestry, AMR: Admixed American ancestry, EAS: East Asian ancestry, SAS: South Asian ancestry

### GIRA is associated with prevalent disease in PMBB

We next assessed the utility of the PGS risk component of the GIRA report by evaluating its ability to identify prevalent disease in PMBB (**Methods**). To validate the PGS performance for eight adult-onset conditions, we associated each condition-specific PGS with its corresponding prevalent condition to find comparable ORs per standard deviation (SD) to those from eMERGE (**Table S6, Methods**). EUR individuals in PMBB had largely similar direction of effects associated with having a score above the specific threshold (‘High Risk-PGS’) compared to the eMERGE reported ORs^55^ across prevalent conditions (**Figure 3; Table S13**). The 95% confidence interval for all prevalent conditions either included or exceeded an OR of 2. CHD individuals in the top 5% of the PGS had higher ORs in PMBB compared to eMERGE (EUR: 3.07 (95% CI: 2.67-3.53) vs 2.36 (95% CI: 2.12-2.62), respectively). In contrast, prevalent CKD had lower ORs across ancestries (e.g. EUR: 1.92 (95% CI: 1.47-2.48) vs 3.41 (95% CI: 3.31-3.52), respectively). HC had lower ORs; however, AFR individuals for HC had higher ORs (AFR PMBB: (4.36 (95% CI: 3.09-6.22)) vs AFR eMERGE (2.98 (95% CI: 1.88-4.60)); **Table S13**). Results were consistent with eMERGE for 5 prevalent conditions in EUR and AFR ancestry when considering overlap in confidence intervals; for example, we observed a 2.5-3.97-fold increased risk for T2D across EUR and AFR individuals compared to 1.76-4.44-fold in eMERGE (**Table S13**). Prior studies also reported that individuals in the top 2% of the genome-wide polygenic score were associated with a 2.66-4.93-fold increased risk for CKD across ancestries^57^ and the top 2% of PGS distribution had a 2.5-4.5-fold increased risk for T2D across ancestries^56,57^. This shows that GIRA’s cross-ancestry PGS along with its ‘High Risk-PGS’ thresholds can identify individuals with increased risk of condition and yield broadly reproducible risk estimates in an independent biobank. However, the variability in ORs for EUR and AFR individuals in PMBB relative to eMERGE reported ORs suggests that the effect of high-risk GIRA may vary across different patient populations to which results will be returned to. Results were also highly variable in non-EUR and AFR populations, likely due to the relatively low sample size and large estimation uncertainties. In addition, differences in sample characteristics and case-control definitions between the eMERGE GIRA study and PMBB may influence predictive performance when applied to cohorts from healthcare systems independent of eMERGE.

**Figure 3.**
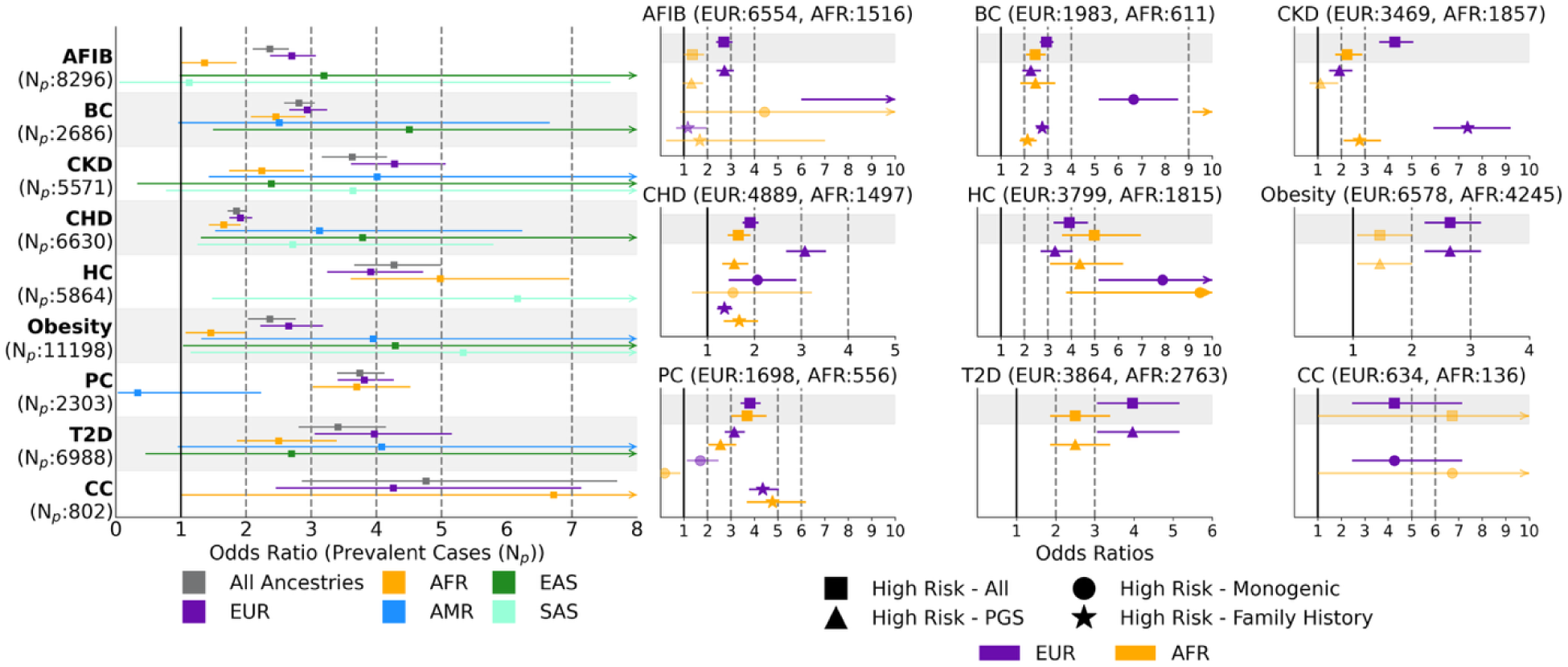
Replication of GIRA results in PMBB across nine prevalent conditions considered by the eMERGE study. PMBB participants were labeled as high-risk for each condition using the eMERGE Genome Informed Risk Assessment (GIRA) together with their condition-specific criteria for each risk indicator. All risk indicators are aggregated into a combined risk score to distinguish individuals at high genetic risk—based on the GIRA-recommended criteria for PGS, monogenic risk, and family history—from those without high genetic risk status. Odds ratios were estimated in a model adjusting for sex, age, age^2^, and first ten genotype or exome PCs. Odds ratios for the combined risk score were stratified by ancestry for prevalent cases (left). Odds ratios for prevalent cases stratified by the three genetic risk components of the GIRA report: Polygenic Risk Score (PGS), Monogenic, and Family History (right). Abbreviations: PMBB: Penn Medicine Biobank, T2D: Type 2 Diabetes, AFIB: Atrial Fibrillation, BC: Breast Cancer, CKD: Chronic Kidney Disease, CC: Colorectal Cancer, CHD: Coronary Heart Disease, HC: Hypercholesterolemia, PC: Prostate Cancer, EUR: European American ancestry, AFR: African/African American ancestry, AMR: Admixed American ancestry, EAS: East Asian ancestry, SAS: South Asian ancestry

For completeness, we also aggregated GIRA high genetic risk components into a binary combined risk score metric, referred to as ‘High Risk - All’. We found ORs ranging from 3.42 (95% CI: 2.81-4.15) for prevalent T2D to 3.63 (95% CI: 3.16-4.17) for prevalent CKD to 4.76 (95% CI: 2.86-7.70) for prevalent CC across ancestries. Except for CHD, all conditions had ORs larger than 2 thus confirming the utility of GIRA as biomarker to identify prevalent cases in a health system biobank. However, AFIB, T2D, CKD, CHD, and obesity showed 95% confidence intervals or ORs that did not reach a threshold of OR > 2 in AFR individuals, highlighting reduced performance in non-EUR populations and underscoring the need for additional ancestry-specific validation for certain conditions (**Figure 3, Table S11**).

As a secondary analysis, we compared the odds ratios for prevalent, before entry into the biobank, conditions to incident conditions (after entry into the biobank) to assess GIRA’s ability to predict incidence cases. Incident case prediction was substantially attenuated for 4 of the 9 conditions – AFIB, CKD, PC, and T2D. This attenuation is consistent with the expectation that incidence risk prediction reflects more conservative effect estimates than prevalent associations as it addresses survival or ascertainment biases. The most significant reductions were observed for CKD (OR before entry: 3.92 (95% CI: 3.26-4.70) vs OR after entry: 1.69 (95% CI: 1.32-2.15)) and AFIB (OR before entry: 2.87 (95% CI: 2.51-3.28) vs OR after entry: 1.42 (95% CI: 1.12-1.79)). For AFIB, this reduction was consistent across all GIRA components: ‘High Risk – PGS’ (OR before entry: 2.87 vs OR after entry: 1.44), ‘High Risk – Monogenic’ (OR before entry: 13.58 vs OR after entry: 5.73), and ‘High Risk – Family History’ (OR before entry: 1.52 vs OR after entry: 0.71). The same was observed for PC. CKD reduction was driven by family history. Thus, GIRA is much weaker at predicting who will newly develop these conditions over time and thus less informative for future disease prediction (Figure S11, Table S18-19).

### GIRA is associated with incident disease in PMBB

Next, we evaluated GIRA’s ability to identify individuals at risk of developing the condition longitudinally focusing on hazard ratios (HRs). We find significant HR across all considered traits albeit with modest magnitudes for T2D, CKD, AFIB, HC and PC (**Figure 4, Table S11**). The lowest signal was for CKD (‘High Risk – All’ HR: 1.82 (95% CI: 1.61-2.05)) and AFIB (‘High Risk – All’ HR: 1.31 (95% CI: 1.05-1.63)). Monogenic component of risk had the strongest association with incident AFIB compared to all other GIRA risk components: ‘High Risk – PGS’ (HR: 1.35 (95% CI: 1.07-1.69)), ‘High Risk – Monogenic’ (HR: 3.46 (95% CI: 1.30-9.24)), and ‘High Risk – Family History’ (HR: 0.57 (95% CI: 0.18-1.77)). For incident CKD, individuals with family history risk were at higher risk of a CKD diagnosis than those with high PGS (HR: 2.46 vs 1.64, respectively). Indeed, across all conditions except AFIB, the 15-year cumulative incidence was substantially higher in individuals at high-risk, highlighting that accounting for time through HRs and cumulative incidence offers a more accurate representation of risk compared to ORs (**Figure 5)**.

**Figure 4.**
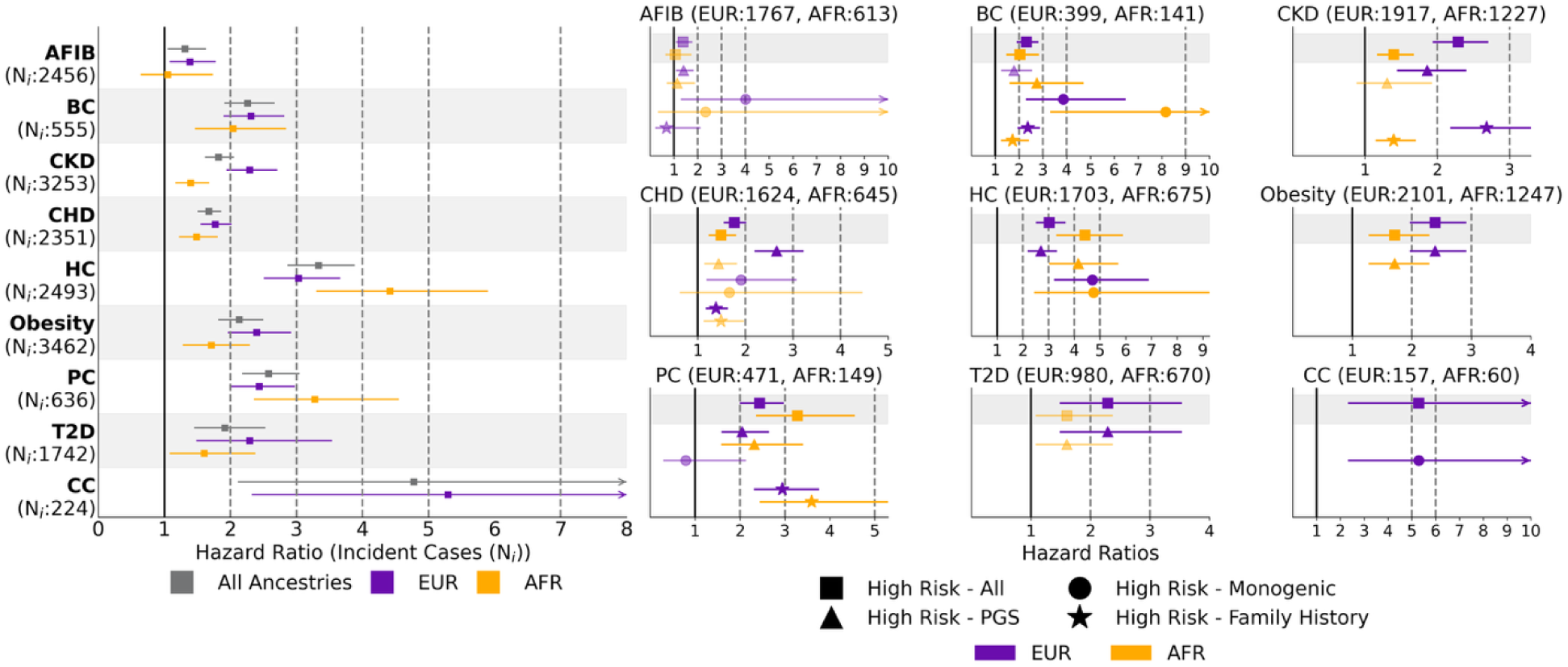
Hazard Ratios for GIRA high risk stratified by the three genetic risk components of the GIRA report: Polygenic Risk Score (PGS), Monogenic, and Family History. High Risk - PGS models were adjusted for sex, age, age^2^, and first ten genotype PCs. High Risk - Monogenic models were adjusted for sex, age, age^2^, and first exome PCs. High Risk - Family History models were adjusted for sex, age, and age^2^. For each model, observation time was defined as the earliest of the following times: date of first diagnosis, last clinical encounter, or death. The disease status was 1 if the individual developed the condition and 0 otherwise. Abbreviations: T2D: Type 2 Diabetes, AFIB: Atrial Fibrillation, BC: Breast Cancer, CKD: Chronic Kidney Disease, CC: Colorectal Cancer, CHD: Coronary Heart Disease, HC: Hypercholesterolemia, PC: Prostate Cancer, EUR: European American ancestry, AFR: African/African American ancestry

**Figure 5.**
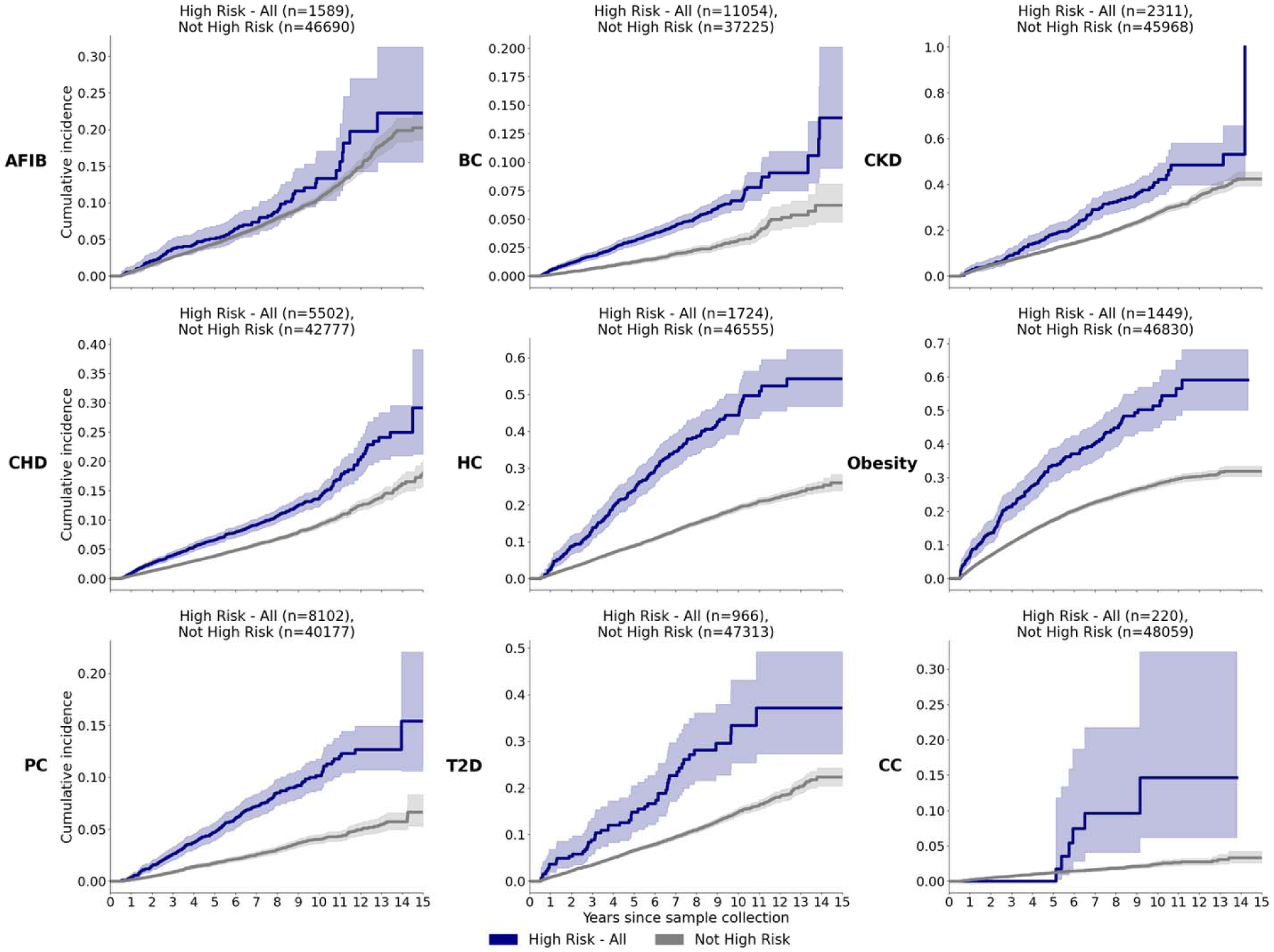
Cumulative incidence curves with stratification by GIRA high-risk versus the rest (‘Not High Risk’). Observation time was defined as the earliest of the following times: date of first diagnosis, last follow-up date within a 15-year follow-up window from sample collection. Participants whose high-risk status for each condition was triggered by one or more of the three GIRA genetic risk factors were aggregated into a ‘High Risk - All’ group. Abbreviations: GIRA: Genome Informed Risk Assessment, T2D: Type 2 Diabetes, AFIB: Atrial Fibrillation, BC: Breast Cancer, CKD: Chronic Kidney Disease, CC: Colorectal Cancer, CHD: Coronary Heart Disease, HC: Hypercholesterolemia, PC: Prostate Cancer

The differential performance of PGS across ancestries is one of the main practical challenges for their use in clinical settings^58,59^. To date, most evaluations of PGS variability across ancestry focused on prevalent disease with limited investigation for incident disease. Stratifying HR across ancestries, we observed effects varying across ancestries for ‘High Risk – All’. Lower HRs were observed for 7 out of 9 in AFR individuals (e.g. T2D EUR: 2.29 (95% CI: 1.48-3.54) vs AFR: 1.60 (95% CI: 1.08-2.38)). However, BC had comparable HRs between EUR and AFR individuals (EUR: 2.31 (95% CI: 1.89-2.82) vs AFR: 2.04 (95% CI: 1.46-2.84)). HC and PC had higher HRs in AFR individuals (e.g. HC EUR: 3.03 (95% CI: 2.51-3.66) vs AFR: 4.41 (95% CI: 3.30-5.90)) (**Table S11, Supplementary Figure 3**).

Finally, we evaluated the 3 components of GIRA to find variability in the predictiveness of the three risk components of GIRA (polygenic, monogenic, family history) across EUR and AFR individuals. For example, EUR individuals with ‘High Risk-Monogenic’ or ‘High Risk-Family History’ for incident BC had higher HRs (‘High Risk-Monogenic’ HR: 3.86 (95% CI: 2.29-6.48); ‘High Risk-Family History’ HR: 2.37 (95% CI: 1.94-2.88)) compared to those with ‘High Risk-PGS’ (HR: 1.79 (95% CI: 1.25-2.55)). The same pattern was observed for HC and AFIB between ‘High Risk-PGS’ and ‘High Risk-Monogenic’. Furthermore, AFR individuals had larger HRs for incident BC for ‘High Risk-PGS’ and ‘High Risk-Monogenic’, but not for ‘High Risk-Family History’ (2.75 (95% CI: 1.60-4.71); 8.16 (95% CI: 3.31-20.1); 1.73 (95% CI: 1.24-2.41), respectively) compared to EUR individuals. However, HRs for incident CHD were lower for ‘High Risk-PGS’ and ‘High Risk-Monogenic’ and higher for ‘High Risk-Family History’ in AFR individuals. Incident PC showed elevated HRs for ‘High Risk-Family History’ in AFR individuals (**Figure 4, Table S11**).

### GIRA-high-risk population demographics differ from general biobank population

Having established the predictive performance of GIRA across the nine adult-onset conditions, we next turned into contextualizing the individuals labeled as high-risk by GIRA. 24,148 (50.1%) of all individuals in PMBB were labeled as high-risk for at least one condition, of which 13,870 (28.7%) due to high PGS risk, 1,111 (2.3%) had monogenic risk, and 13,873 (28.8%) reported a family history of disease. 14,676 (30.4%) were at high-risk due to the polygenic and/or monogenic risk components larger than the eMERGE expected results (25%^58^). This shows GIRA’s potential to identify a larger proportion of the population that could benefit from early intervention while also highlighting the disruptive impact and burden on the health system if GIRA is implemented in clinical practice. Among high-risk individuals, the proportion of those who “already have disease” ranged from 2.5% for PC to 11.0% for obesity. More specifically, 2.5% of the 8,102 individuals at high PC risk, 12.9% of 2,311 at high CKD risk, and 11.0% of 1,449 at high obesity risk developed the corresponding condition during follow-up. These relatively low percentages suggest that most high-risk individuals have not yet manifested clinically recognizable disease within the observation time, which aligns with risk assessments ability to identify risk before disease onset^60^. Of the individuals labeled as high-risk by GIRA’s criteria across all conditions, 5,023 (20.8%) individuals developed at least one condition during follow-up. Lastly, there were 6,965 PMBB individuals who developed at least one condition during follow-up but were not at high-risk. Thus, these findings indicate that case proportions among high-risk individuals likely underestimate the true burden of disease due to these individuals having not yet developed the disease or remain unclassified due to phenotyping uncertainty.

Next, to understand variability in high-risk rates in PMBB subpopulations, we compared the high-risk proportion among individuals in different ancestries and social deprivation index quartiles to those for all PMBB individuals. We found significantly higher high-risk rates among individuals of AFR ancestry (High Risk – All: 56.6% vs 50.1%, Z=-12.5, p=7.43×10^-36^, respectively), largely driven by high PGS and family history. Higher rates in AFR individuals were not driven by monogenic risk, which saw lower high-risk rates compared to all high monogenic risk individuals (1.6% vs 2.3%, Z=4.77, p=1.85×10^-6^, respectively). In contrast, lower-risk rates were observed for EUR (driven by high PGS and family history), EAS (driven solely by family history), and SAS individuals (e.g. SAS High Risk - All: 40.0% vs 50.1%, Z = 5.25, p = 1.49×10^-7^). Notably, the lower rates in East Asian ancestries being driven solely by family history indicated a large impact from factors like environment, recall bias, or reporting behavior than underlying genetic risk (**Table S15-16**). Second, to understand how socioeconomic factors may impact GIRA’s utility, we determined high-risk proportions among social deprivation index (SDI) quartiles. Compared to all high-risk PMBB individuals, there were significantly higher high-risk rates among the most deprived (SDI quartile 4) (53.7% vs 50.1%, Z=-6.77, p=1.30×10^-11^, respectively), which were driven by high PGS risk and high family history risk. Additionally, lower high PGS risk rates were observed in the least deprived quartile (SDI quartile 1). Monogenic risk, however, had lower high-risk rates at SDI quartile 4 (1.7% vs. 2.3%, Z = 3.98, p=6.79×10^-5^) (**Table S17**), suggesting the GIRA’s polygenic risk and family history performance is more impacted by lower socioeconomic status than monogenic risk as illustrated in previous studies^61,62^. Lastly, we evaluated the demographic composition of the GIRA high-risk group versus the incident case population across the nine conditions. Except obesity, CKD, and AFIB, ancestry enrichments among high-risk individuals largely mirrored those observed in incidence rates. Specifically, obesity showed no enrichment for EUR or AFR individuals among high-risk individuals, whereas incident obesity was overrepresented in AFR individuals and underrepresented in EUR individuals. High-risk individuals for CKD were enriched for EUR and depleted for AFR individuals, while the opposite pattern was seen for incident CKD (**Figure 7**). This under-enrichment of AFR individuals within the GIRA-classified high-risk individuals may be due to the underperformance of genetic risk in AFR individuals relative to EUR individuals^63^. As a secondary analysis, we restricted to individuals not lost during follow-up (**Methods**) to find our results remained largely consistent (**Supplementary Figures 8-10**).

**Figure 6.**
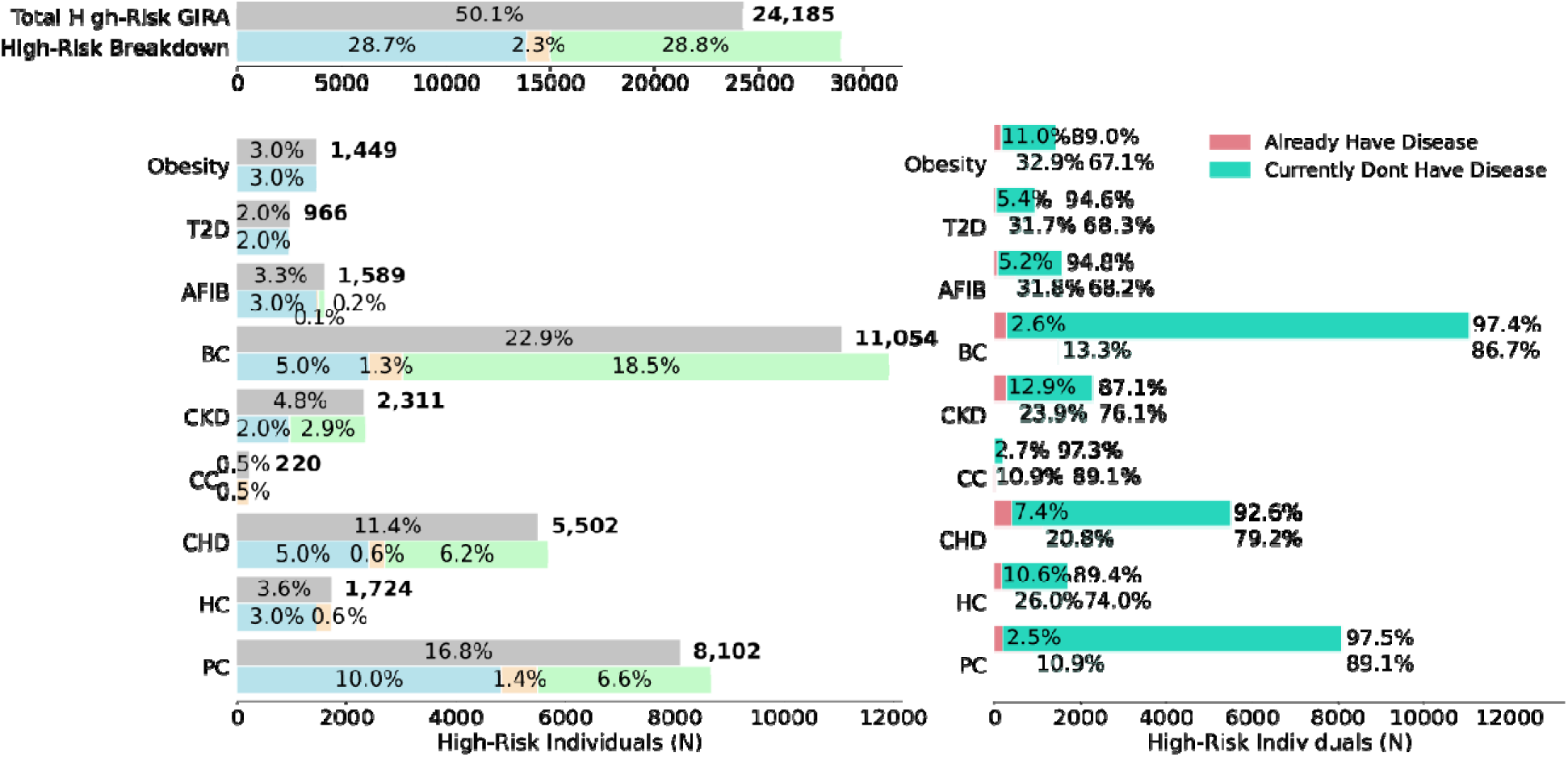
Characterization of the PMBB participants labeled as High-Risk using GIRA guidelines for nine conditions. The left panel shows the percentage of unique PMBB participants classified as high-risk based on one or more of the three genetic risk components in the GIRA report. Grey bars indicate the overall proportion of high-risk individuals across all and within each condition, while colored bars show the contribution of each specific component–PGS, monogenic risk, or family history–to high-risk status. The right panel shows the disease status of each high-risk individual across conditions, categorized as case (“Already Have Disease”) or the remaining PMBB participants (“Currently Don’t Have Disease”). Prevalent cases (lighter bar) are compared to incident cases (darker bar). Abbreviations: PMBB: Penn Medicine Biobank, GIRA: Genome Informed Risk Assessment, T2D: Type 2 Diabetes, AFIB: Atrial Fibrillation, BC: Breast Cancer, CKD: Chronic Kidney Disease, CC: Colorectal Cancer, CHD: Coronary Heart Disease, HC: Hypercholesterolemia, PC: Prostate Cancer

**Figure 7.**
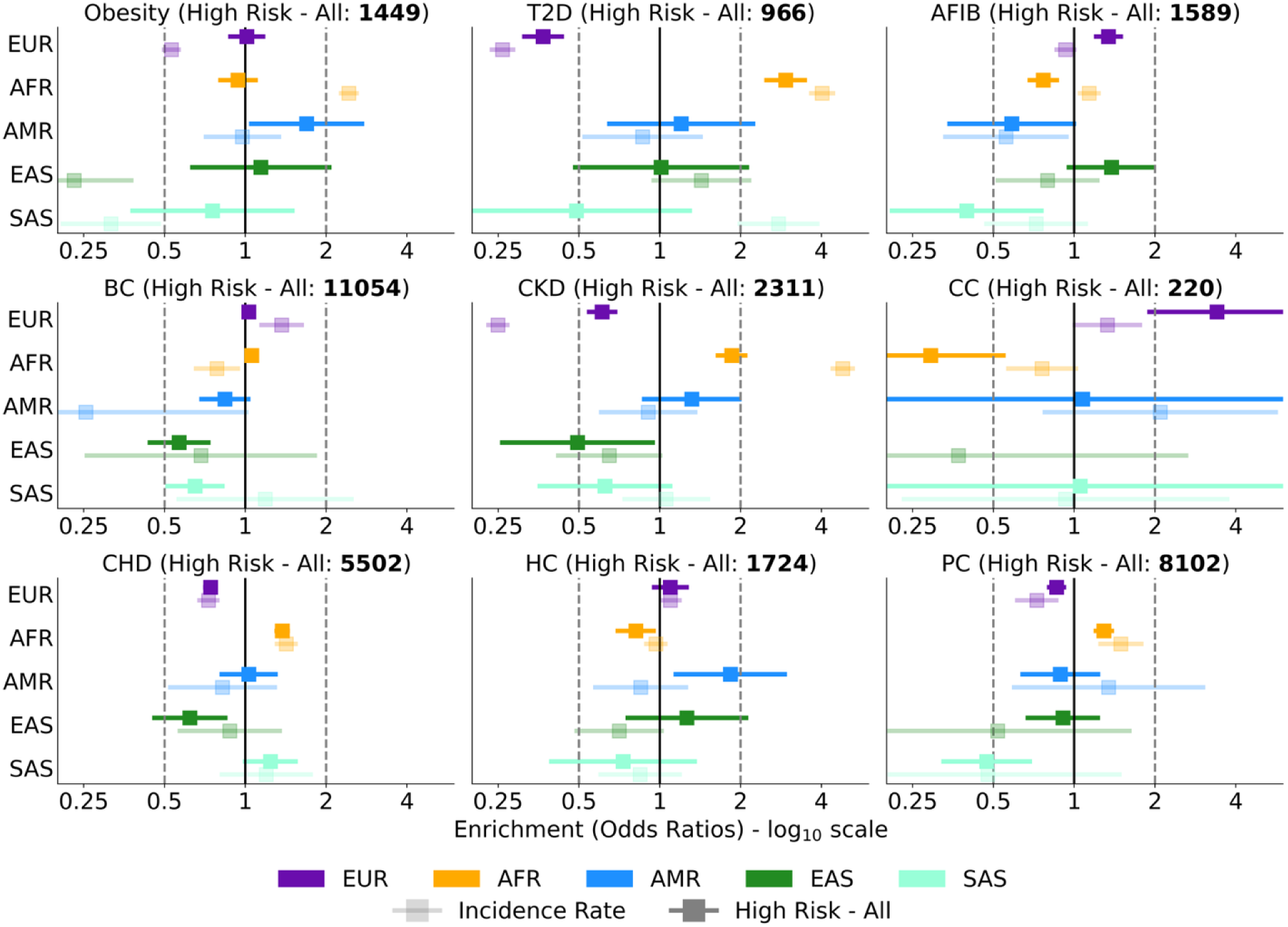
Ancestry enrichments among high-risk PMBB participants across nine conditions compared with overall PMBB enrichments. Enrichment within incident (lighter shade) cases versus PMBB participants labeled as high-risk (darker shade) across the nine adult GIRA conditions is estimated using odds ratios and their standard errors. High Risk – All error bar for CC in EAS ancestry individuals is not plotted due to small sample size. Odds ratios are plotted on the log_10_ scale. Cutoff bars extend beyond the displayed range and were truncated at x-axis limits of 0.25 and 6 for visualization. Abbreviations: PMBB: Penn Medicine Biobank, GIRA: Genome Informed Risk Assessment, T2D: Type 2 Diabetes, AFIB: Atrial Fibrillation, BC: Breast Cancer, CKD: Chronic Kidney Disease, CC: Colorectal Cancer, CHD: Coronary Heart Disease, HC: Hypercholesterolemia, PC: Prostate Cancer, EUR: European American ancestry, AFR: African/African American ancestry, AMR: Admixed American ancestry, EAS: East Asian ancestry, SAS: South Asian ancestry

## Discussion

The Penn Medicine Biobank as institutional biobank embedded within the University of Pennsylvania’s Healthcare System (UPHS) provided an ideal setting for evaluation of the condition-specific high-risk GIRA criteria, informed by eMERGE, in a real-world health system. We showed that GIRA high PGS can identify individuals with a 2.81 to 3.41-fold increased risk depending on the prevalent condition, demonstrating the PGS in this analysis largely aligned with the Lennon et al eMERGE reported results. The clinical translation of the high-risk criteria in GIRA relies on its ability to predict future disease cases rather than the strength of the association with prevalent conditions. We presented the first study that evaluated GIRA’s high-risk criteria longitudinally showing that hazards ratios show modest associations with each GIRA risk component for various conditions. We demonstrated differences in high-risk classification across ancestry and social deprivation index quartiles. Finally, we report that GIRA high-risk individuals differed in their demographic composition compared to incident cases suggesting that additional factors, such as social determinants of health or environmental exposures, may contribute to disease development beyond genetic risk. This was further stressed by lower high-risk rates in East and South Asian ancestries being driven by family history and higher high-risk rates in the most deprived social deprivation index quartile for high PGS and family history. Our analysis demonstrates that though GIRA’s high-risk criteria is largely generalizable for prevalent conditions, real-world implementation will require careful choice of metrics to convey magnitude of risk, potential condition-specific calibration, and ancestry-stratified performance metrics to accurately communicate risk across diverse populations.

Half of PMBB participants were identified as high-risk for at least one condition, highlighting the substantial burden that returning GIRA results could place on the healthcare system. 28.7% of PMBB participants had at least one high-risk PGS result, which is greater than the 23.2%^62,63^ in the first 2,500 processed samples in the GIRA study. However, the Genomic Medicine at Veterans Affairs (GenoVA) study, which is providing insights for evaluating the clinical utility of monogenic and PGS implementation for six common diseases in primary care settings, estimated an expected 33% at high-risk PGS^64,65^. More specifically, in their first 840 processed samples, 2% were at high-risk monogenic while 37.1% had at least one high-risk PGS result. We also observed 30.4% PMBB individuals with a high-risk PGS or monogenic result while eMERGE expected 25%. The proportion of high-PGS risk individuals in PMBB fell in between those reported by the GIRA and GenoVa studies, while the 2.3% for high-risk monogenic closely mirrored the reported 2% by the GenoVa study suggesting that the risk assessments are largely generalizable and that differences in high-risk proportions among different studies likely reflect differences in cohort composition, PGS thresholds, and the specific PGS implemented, rather than differences in underlying genetic risk. It is important to note that eMERGE determined high-risk results for breast cancer using BOADICEA; however, our analysis focused only on the high PGS for breast cancer as not all components needed for the integrated risk score were available in PMBB.

We conclude with several limitations of our work. First, even with current efforts to enroll a diverse group of individuals into PMBB, the relatively small sample size for Admixed American, East Asian, and South Asian individuals caused large estimation uncertainties. Even with differences in the proportion of diverse individuals, we observed OR > 2 for EUR and AFR individuals in PMBB; however, the increase in risk varied by population as compared to eMERGE, likely reflecting differential performance of these score to predict disease risk across health care systems with different sample characteristics^11,66^. Validating GIRA in a cohort with different ancestry proportions remains valuable, as it evaluates its performance and potential biases in a setting more representative of real-world healthcare systems. PMBB provided such a setting as the biobank is embedded within the UPHS. More specifically, PMBB participants have comparable age and gender distributions, but a higher proportion of individuals who self-identify as African American (24.6% vs 18.2%) or White (66.8% vs 55.0%) compared with the University of Pennsylvania Healthcare System^23^. Second, PMBB is composed of individuals recruited from the UPHS, a population potentially exhibiting a higher disease burden than the general population. This inflates prevalence estimates and monogenic variant penetrance, which carries bias and may limit the generalizability of these findings. Third, lack of precision in family history information in the EHR due to recall bias and sensitivity to wording in queries may lead to inaccurate risk assessment^67,68^. Additionally, while eMERGE used the MeTree family history tool to collect detailed information on first- and second-degree relatives directly from participants^1^, PMBB relied on available relative information, which served as a reasonable proxy. Fourth, because the case-control definitions used by eMERGE were not available, we instead applied validated EHR-based phenotyping algorithms with high positive predictive value^69^. The resulting prevalence estimates in PMBB generally fell between those reported in population-based studies and those observed in eMERGE, supporting the validity of our phenotyping approach. Fifth, direct comparison of logistic regression-derived ORs with cox regression-derived HRs is limited by the differences in what they measure as HRs compares instantaneous event rates over follow-up and ORs compares odds of being a case at a fixed timepoint. Due to this, we compared ORs for prevalent (before entry) cases to incident cases to find significant attenuation in effect sizes for 4 out of 9 conditions suggesting GIRAs limited utility for future disease prediction. In addition, ORs may be impacted by survivorship bias in prevalent cases which may attenuate the association. Lastly, incidence disease prediction may be impacted by variability in follow-up times, phenotyping strategies, and recruitment biases in a biobank population.

## Methods

### Study population

The Penn Medicine Biobank (PMBB) is an electronic health record linked biobank at the University of Pennsylvania that integrates an academic health system that serves south-central Pennsylvania and New Jersey as well as northern Delaware with genomic and exome data^23^. PMBB agnostically recruits individuals from outpatient clinical settings from whom it obtains consent to their EHR data as well as to collection and storage of biospecimens and genetic sequencing data. These participants underwent genome and exome sequencing at Regeneron Genetics Center. Clinical information from 57,170 sequenced individuals, of which approximately 26.3% of participants are of non-European ancestry, makes PMBB one of the more diverse medical biobanks. Clinical diagnosis, procedural, visit, laboratory, prescription, and demographic information was used to define each condition.

### Genotype quality control and imputation

Detailed information on genotyping and imputation for Freeze 2.0 can be found in a previous publication^23^. Briefly, in Freeze 2.0, 43,913 PMBB participants were genotyped using a custom genotyping array constructed from the Global Screening Array. An additional 13,257 individuals were genotyped via genotype by sequencing capture as part of Freeze 3.0. To ensure data quality, SNPs with more than 5% missingness were excluded. Samples with over 10% missingness or sex discrepancies were also removed using PLINK 2.0. Following genotype quality control, imputation was conducted using the TOPMed version R3 reference panel via the Michigan TOPMed Imputation Server 2 with minimac v4.1.6. Phasing was carried out using Eagle v2.4 and the GRCh38/hg38 genome build. The data were imputed in separate batches, so the final quality control step involved merging all batches into a single dataset. An OR logic was then applied to filter variants based on the following criteria: an average R² greater than 0.2, sequenced in Freeze 2.0, or sequenced in Freeze 3.0.

### Exome sequencing and quality control

Whole exome sequencing (WES) on 57,170 individuals in the Penn Medicine Biobank was performed at the Regeneron Genetics Center (Tarrytown, NY) as described previously^23^. This total consists of 43,913 samples from Freeze 2.0 and an additional 13,257 sequenced in 2024 as part of Freeze 3.0. DNA libraries were prepared using a custom NEDNext Ultra II FS DNA library prep kit to generate fragments at an average size of 200 base pairs. During library amplification with KAPA HiFi polymerase, a Y-shaped adapter was ligated and asymmetric 10 base pair barcodes were added to enable exome capture and sequencing. DNA samples in Freeze 2.0 were processed for exome capture on a custom IDT xGen probe library. Freeze 3.0 used the RGC-developed Twist Comprehensive Exome panel for exome capture. Samples from both freezes were then sequenced on an Illumina NovaSeq 6000 system using S4 flow cells.

Sequencing reads were aligned to the human reference genome build GRCh38 using the Burrows-Wheeler Aligner - MEM algorithm. To ensure consistency given the differences in exome capture products, the datasets were reprocessed and jointly recalled across the combined set of their respective calling regions. Variants were jointly called using GLnexus v1.4.3 with the DeepVariantWES configuration. Samples with contamination more than 5%, less than 80% of target bases at 20x coverage, gender discordance, plating errors, duplicates, and no genotyping array data were removed.

### Population stratification

To infer genetically defined ancestry groups, principal component analysis (PCA) was performed using the *smartpca* module from the EIGENSOFT package. PCA was conducted on a linkage disequilibrium (LD)-pruned set of 182,701 common SNPs that had undergone quality control, including removal of SNPs that failed Hardy-Weinberg equilibrium test (p < 1e-6), had a minor allele frequency below 0.1, and related individuals. The study population was assigned to genetically inferred ancestry (GIA) groups - European American (EUR), Admixed American (AMR), African/African American (AFR), East Asian (EAS), and South Asian (SAS) – using quantitative discriminant analysis (QDA) based on the top 20 principal components. The continental ancestry populations from 1000 Genomes Project were used as a reference. QDA assigned each PMBB individual to the closest ancestry group based on genetic similarity in principal component space.

For ancestry-specific analysis, participants with complete information on the defined condition and covariates were stratified by European American and African American ancestry groups, which had sufficient sample sizes for analysis.

### Phenotype definition

We adapted validated electronic phenotype algorithms from the Phenotype KnowledgeBase (PheKB) to define case and control status for four conditions – Breast Cancer, Hypercholesterolemia, Colorectal Cancer, and Chronic Kidney Disease. PheKb is a collaborative online environment created by the eMERGE network to serve as a central repository for the development, shareability, and validity of algorithms using health care generated data across implementation sites.^69^ Details on the phenotyping algorithms are provided in the **Supplementary Methods**. We adapted the chronic kidney disease algorithm to focus on cases that had an estimated glomerular filtration rate (eGFR) below 60 ml/min/1.73m2 or received a dialysis or kidney transplant without the incorporation of staging CKD based on albuminuria or eGFR.^63,70^

Atrial fibrillation (AFIB) algorithm was adapted from ICD-9 and 10 diagnosis code-based case definition outlined in the Chronic Conditions Data Warehouse (CCW). The CCW algorithms were developed from claims data using research literature, expert panels, and criteria used by other federal sources and have been previously applied as phenotyping methods to EHR data to ascertain chronic disease status.^71^ For an individual to be classified as having AFIB based on the CCW algorithm, they must have at least 1 inpatient claim or 2 hospital outpatient claims with diagnosis codes that are at least 30 days apart but within 1 year. A previous study found 1 inpatient or 2 outpatient codes separated by >30 days but within 1 year had a PPV of 78.7% and sensitivity of 92.5% in their prevalent approach while their incident approach had a PVV of 69.7% and sensitivity of 93.6%.^72^

Coronary Heart Disease, Prostate Cancer, and Obesity algorithms were adapted from the Polygenic Risk Methods in Diverse Populations (PRIMED) Consortium. Lastly, we implemented the most up to date phenotyping algorithm for type 2 diabetes from the Szczerbinski et al. study^73^. This study developed and validated a phenotyping algorithm for type 2 diabetes, modified from the Northwestern University algorithm in PheKB^74^, in the All of Us cohort using electronic health records as well as a universal algorithm to identify individuals without diabetes.^73^

Case or control status for breast and prostate cancer were assigned to only females and males, respectively. For each of the conditions, we considered the earliest date at which the diagnosis was ascertained as the diagnosis date. All participants diagnosed at dates prior to, at, or post sample collection (baseline) in PMBB were considered prevalent, while participants diagnosed after sample collection were considered incident. Additional details on the ascertainment of each condition are provided in the **Supplementary Methods**.

### Monogenic risk score classification

Genetic variants were annotated using Ensembl’s Variant Effect Predictor (VEP) version 113.0^75^ to access population allele frequencies from the gnomAD Genome Aggregation Database, clinical significance as reported by ClinVar, and predicted gene impact. The analysis focused on rare, high-quality variants located within protein-coding regions of 16 genes associated with the six monogenic conditions included in the GIRA study: atrial fibrillation (*LMNA*); breast cancer (*BRCA1, BRCA2, PALB2, PTEN, TP53, STK11*); colorectal cancer (*EPCAM, MLH1, MSH2, MSH6, PMS2, STK11, PTEN, TP53*); coronary heart disease (*APOB, LDLR, LDLRAP1, PCSK9*); hypercholesterolemia (*APOB, LDLR, LDLRAP1, PCSK9*); and prostate cancer (*BRCA1, BRCA2, EPCAM, MLH1, MSH2, MSH6, PMS2*). Variants with a minor allele frequency (MAF) greater than 0.005 in any of the following gnomAD racial subpopulations—Non-Finnish European, African, or Latino—were excluded from the analysis as well as synonymous variants. We did not consider superpopulation maximum allele frequencies. We further selected variants classified in ClinVar with designations indicating pathogenicity, including *Pathogenic*, *Pathogenic/Likely pathogenic*, *Likely pathogenic*, *Pathogenic/Likely pathogenic/Pathogenic, low penetrance*, *Pathogenic/Likely pathogenic | risk factor*, *Pathogenic | association | protective*, *Likely pathogenic | risk factor*, *Pathogenic/Likely pathogenic | other*, and *Pathogenic | other*. Only variants submitted by multiple contributors or reviewed by an expert panel were retained.

### Polygenic risk score derivation and ancestry calibration

To ensure our validation of GIRA’s high-risk criteria in PMBB closely mirrored the original implementation decisions made in the eMERGE study, we used the same polygenic risk scores that were employed in their study. Each condition’s PGS was calculated by multiplying individual risk allele dosages by their corresponding weights that were downloaded from the eMERGE GitHub repository (https://github.com/broadinstitute/eMERGE-implemented-PRS-models-Lennon-et-al) using pgsc_calc. These scores are also available on the UCSC Genome Browser (https://genome.ucsc.edu/s/Max/emerge), and PGS Catalog IDs for most scores are detailed in the Lennon et al. publication^29^. As the SNP weights used for obesity in the GIRA study were not publicly available, we evaluated the performance of all published obesity PGS from the PGS Catalog within the All of Us Research Program (AoU) to identify the best-performing score^30^. Within all participants of AoU v7, we applied each published obesity PGS, then standardized and residualized out ancestry PCs from each PGS as described previously^76^. We then estimated the area under the receiver operator curve (AUC) for logistic regression models when predicting obesity status based on the PGS, adjusting for age, sex, 10 PCs, and sequencing site. The obesity PGS with the highest AUC was selected for downstream analyses within PMBB.

**Supplementary Table 14** reports the number of single nucleotide polymorphisms (SNPs) in the base file for each published PGS and the proportion available in imputed PMBB data, showing ≥75% overlap depending on the PGS. Performance of each PGS in PMBB was consistent with the odds ratios per standard deviation reported by eMERGE.

To achieve comparable score distributions across ancestry groups, the raw PGSs were calibrated using genetic principal components, implemented through the –run_ancestry method in pgsc_calc. In brief, this adjustment accounts for differences in the mean and variance of PGS distributions between ancestries, following the calibration of ancestry for PGS approach used in the eMERGE Network’s GIRA report. As a result, similar proportions of individuals in each ancestry group fall above a given percentile threshold. For each condition, the GIRA report defined ancestry-independent thresholds above which individuals were classified as “high risk.” These thresholds differed by condition. After calibrating the PGS for ancestry, scores were converted into a categorical variable to distinguish individuals at high polygenic risk, based on the GIRA-recommended percentile cutoff, from those below the threshold.

### Family history classification

Family history (FamHx) information from the EHR was used to identify individuals with a family history for five GIRA conditions where family history is considered a risk factor. To identify FamHx of specific conditions, we extracted the following FamHx information:

- Atrial fibrillation: Individuals whose parents had a history of ‘Atrial fibrillation’
- Breast cancer: Individuals whose first and second-degree relatives had a history of ‘breast cancer’, ‘Bilateral breast cancer’, ‘Postmenopausal breast cancer’, ‘Premenopausal breast cancer’, ‘Left Breast Cancer’, and ‘Right Breast Cancer
- Chronic kidney disease: Individuals whose first-degree relatives had a history of ‘Kidney Disease’, ‘Kidney Disorder’, ‘Chronic Kidney Disease’, and ‘Polycystic Kidney Disease’
- Coronary heart disease: Individuals whose first-degree relatives had a history of ‘Heart Disease’
- Prostate cancer: Individuals whose first-degree male relatives had a history of ‘Prostate Cancer’

First-degree relatives included an individual’s parents, siblings, and children. Second-degree relatives included an individual’s grandparents, grandchildren, uncles, aunts, nephews, nieces, and half-siblings.

Monogenic risk and family history components of the GIRA report could not be validated, as eMERGE did not release these corresponding results.

### Statistical Analysis

All analysis used Python 3 or R 4.2.1.

To align our population with the eMERGE GIRA study, we restricted our analysis to adults aged 18 to 75 at enrollment. However, eMERGE-defined age bands of return were not incorporated in our retrospective analysis when defining whether GIRA was ‘high-risk’ or actionable. Although the primary objective was to predict incident disease events, prevalent conditions were included in the initial analyses to enable comparison with the findings of Lennon et al, who reported odds ratios associated with having a score above a specified threshold^18^. To validate the high-PGS result in Lennon et al for each prevalent condition, logistic regression models, both across and stratified by ancestry, were used to estimate associations with high-risk status for each condition, adjusting for the first ten genotype or exome principal components, age, age², and sex. Models were also run separately for each GIRA risk component, PGS, monogenic risk, and family history, as well as for a combined risk score, labeled as ‘High Risk - All’, aggregating all high-risk indicators to distinguish individuals at high genetic risk from those without. We focused on the odds ratios for each high-risk component in EUR and AFR ancestry groups, given the limited sample sizes of other ancestry groups across conditions. As a secondary analysis, we compared odds ratios for prevalent before entry into biobank cases with incident cases, stratified by each GIRA component.

Subsequent analyses focused on only incident disease cases. Cox proportional hazard models were performed for each condition to estimate hazard ratios comparing those at high-risk based on each GIRA risk component or the combined risk score with those not at high risk. We report hazard ratios for EUR and AFR groups for each high-risk component. For each hazard ratio model, observation time was defined as the earliest of the following times: date of first diagnosis, last clinical encounter, or death. The disease status was 1 if the individual developed the condition and 0 otherwise. We estimate time-dependent disease incidences for each condition stratified by high-risk status versus those not at high-risk using 1 minus the Kaplan-Meier estimator. Observation time was defined as the earliest of the following times: date of first diagnosis, last follow-up date within a 15-year follow-up window from the time of sample collection, or death. High-risk status was assigned to participants with at least one of the three GIRA genetic risk indicators.

Odds ratios and 95% confidence intervals for ancestry, age, or sex enrichment were estimated using logistic regression models comparing individuals in one context group to those in all the other context groups for both prevalent and incidence cases. Ancestry enrichment models were adjusted for age, age², sex, and batch. Age enrichment models were adjusted for sex and batch. Sex enrichment models were adjusted for age, age², and batch. The same models were repeated to assess enrichment among high-risk PMBB participants across nine conditions. Since we only consider females or males for breast cancer and prostate cancer, respectively, we don’t estimate the odds ratios for sex enrichment.

To compare high-risk proportions across ancestry strata and social deprivation index (SDI) quartiles with those observed in the overall PMBB population, we performed two-proportion z-tests to evaluate whether high-risk percentages in specific subpopulations differed significantly from the overall high-risk percentage. Ancestry-specific high-risk proportions were calculated by determining the proportion classified as ‘High Risk – All’ and the high-risk proportion for each genetic risk component within each ancestry stratum. P-values were adjusted for multiple testing across 24 comparisons (four high-risk categories across six ancestry strata). Ancestry enrichment for each high-risk categories was also estimated using logistic regression models comparing individuals in one ancestry strata to all other ancestries (**Table S16**). The models were adjusted for age, age², sex, and batch. SDI scores were categorized into quartiles. High-risk proportions within each SDI quartile were calculated using the same approach as for ancestry strata. P values were adjusted for multiple testing across 16 comparisons (four high-risk categories across four SDI quartiles).

In secondary analysis, we excluded individuals who were lost during follow-up. Individuals not lost during follow-up were defined as those who had at least one encounter in the two years prior to the last encounter date.

## Data Availability

Researchers can access the Penn Medicine Biobank individual-level genotype, exome, and phenotype data following its data application procedures.

## Acknowledgements

We acknowledge the Penn Medicine BioBank (PMBB) for providing data and thank the patient-participants of Penn Medicine who consented to participate in this research program. We would also like to thank the Penn Medicine BioBank team and Regeneron Genetics Center for providing genetic variant data for analysis. The PMBB is approved under IRB protocol# 813913 and supported by Perelman School of Medicine at University of Pennsylvania, a gift from the Smilow family, and the National Center for Advancing Translational Sciences of the National Institutes of Health under CTSA award number UL1TR001878.

We thank Rex L. Chisholm, Noura Abul-Husn, Jordan W. Smoller, Josh F. Peterson, Richard Sharp, Eimear Kenny, Cynthia C. Williams, Christina Alejandra Rodrigues, and Leah Kottyan for their feedback on this manuscript.

## References

1. Returning integrated genomic risk and clinical recommendations: The eMERGE study. Genetics in Medicine 25, 100006 (2023).

2. Daly, M. B. et al. NCCN Guidelines Insights: Genetic/Familial High-Risk Assessment: Breast, ovarian, and pancreatic, version 1.2020. J. Natl. Compr. Canc. Netw. 18, 380–391 (2020).

3. Gupta, S. et al. NCCN Guidelines Insights: Genetic/Familial High-Risk Assessment: Colorectal, version 3.2017. J. Natl. Compr. Canc. Netw. 15, 1465–1475 (2017).

4. Khera, A. V. et al. Genome-wide polygenic scores for common diseases identify individuals with risk equivalent to monogenic mutations. Nature Genetics 50, 1219–1224 (2018).

5. Abul-Husn, N. S. et al. Genetic identification of familial hypercholesterolemia within a single U.S. health care system. Science 354, aaf7000 (2016).

6. Mars, N. et al. Systematic comparison of family history and polygenic risk across 24 common diseases. American journal of human genetics 109, (2022).

7. Lu, T., Forgetta, V., Richards, J. B. & Greenwood, C. M. T. Capturing additional genetic risk from family history for improved polygenic risk prediction. *Commun*. Biol. 5, 595 (2022).

8. Hujoel, M. L. A., Gazal, S., Loh, P.-R., Patterson, N. & Price, A. L. Liability threshold modeling of case-control status and family history of disease increases association power. Nat. Genet. 52, 541–547 (2020).

9. Guttmacher, A. E., Collins, F. S. & Carmona, R. H. The family history--more important than ever. N. Engl. J. Med. 351, 2333–2336 (2004).

10. Valdez, R., Yoon, P. W., Qureshi, N., Green, R. F. & Khoury, M. J. Family history in public health practice: a genomic tool for disease prevention and health promotion. Annu. Rev. Public Health 31, 69–87 1 p following 87 (2010).

11. Conti, D. V. et al. Trans-ancestry genome-wide association meta-analysis of prostate cancer identifies new susceptibility loci and informs genetic risk prediction. Nat. Genet. 53, 65–75 (2021).

12. Ge, T. et al. Development and validation of a trans-ancestry polygenic risk score for type 2 diabetes in diverse populations. Genome Med. 14, 70 (2022).

13. Fahed, A. C. et al. Polygenic background modifies penetrance of monogenic variants for tier 1 genomic conditions. Nat Commun 11, 3635 (2020).

14. Murray Leech, J., et al. Common genetic variants modify disease risk and clinical presentation in monogenic diabetes. Nat. Metab. 7, 1819–1829 (2025).

15. Saadatagah, S. et al. Polygenic Risk, Rare Variants, and Family History: Independent and Additive Effects on Coronary Heart Disease. JACC Adv 2, 100567 (2023).

16. McCarty, C. A. et al. The eMERGE Network: A consortium of biorepositories linked to electronic medical records data for conducting genomic studies. BMC Medical Genomics 4, 1–11 (2011).

17. The Electronic Medical Records and Genomics (eMERGE) Network: past, present, and future. Genetics in Medicine 15, 761–771 (2013).

18. Lennon, N. J. et al. Selection, optimization and validation of ten chronic disease polygenic risk scores for clinical implementation in diverse US populations. Nature Medicine 30, 480– 487 (2024).

19. Kullo, I. J. Clinical use of polygenic risk scores: current status, barriers and future directions. Nature Reviews Genetics 1–18 (2025).

20. Limdi, N. et al. The Electronic Medical Records and Genomics study: Design and analytic framework for assessing the impact of genome-informed risk assessments. Am. J. Hum. Genet. 113, 664–677 (2026).

21. Collins, G. S., Reitsma, J. B., Altman, D. G. & Moons, K. G. M. Transparent Reporting of a multivariable prediction model for Individual Prognosis or Diagnosis (TRIPOD): the TRIPOD statement. Ann. Intern. Med. 162, 55–63 (2015).

22. Xiang, R. et al. Recent advances in polygenic scores: translation, equitability, methods and FAIR tools. Genome Med. 16, 33 (2024).

23. Verma, A. et al. The Penn Medicine BioBank: Towards a Genomics-Enabled Learning Healthcare System to Accelerate Precision Medicine in a Diverse Population. Journal of Personalized Medicine 12, 1974 (2022).

24. Pfeiffer, R. M. & Gail, M. H. Two criteria for evaluating risk prediction models. Biometrics 67, 1057–1065 (2011).

25. McHugh, J. K. et al. Assessment of a polygenic risk score in screening for prostate cancer. N. Engl. J. Med. 392, 1406–1417 (2025).

26. Chatterjee, N., Shi, J. & García-Closas, M. Developing and evaluating polygenic risk prediction models for stratified disease prevention. Nat. Rev. Genet. 17, 392–406 (2016).

27. Kraft, P. et al. Beyond odds ratios--communicating disease risk based on genetic profiles. Nat. Rev. Genet. 10, 264–269 (2009).

28. Elliott, J. et al. Predictive accuracy of a polygenic risk score-enhanced prediction model vs a clinical risk score for coronary artery disease. JAMA 323, 636–645 (2020).

29. Haas, R. et al. Diverse genomes, shared health: Insights from a health system biobank. medRxiv (2025) doi:10.1101/2025.06.11.25329386.

30. All of Us Research Program Genomics Investigators. Genomic data in the All of Us Research Program. Nature 627, 340–346 (2024).

31. Piekos, J. A. et al. Evaluating the relationships between genetic ancestry and the clinical phenome. Pac. Symp. Biocomput. 29, 389–403 (2024).

32. Heart disease prevalence. https://www.cdc.gov/nchs/hus/topics/heart-disease-prevalence.htm (2024).

33. Khan, M. A. B. et al. Epidemiology of type 2 diabetes - Global Burden of Disease and forecasted trends. J. Epidemiol. Glob. Health 10, 107–111 (2020).

34. https://jamanetwork.com/journals/jamanetworkopen/fullarticle/2801355.

35. CDC. Chronic kidney disease in the United States, 2023. Chronic Kidney Disease https://www.cdc.gov/kidney-disease/php/data-research/index.html (2025).

36. van Alten, S., Domingue, B. W., Faul, J., Galama, T. & Marees, A. T. Reweighting UK Biobank corrects for pervasive selection bias due to volunteering. Int. J. Epidemiol. 53, (2024).

37. Pimplaskar, A. et al. Inclusion bias affects common variant discovery and replication in a health-system linked biobank. medRxiv (2025) doi:10.1101/2025.04.04.25325131.

38. Cancer of the breast (female) - cancer stat facts. SEER https://seer.cancer.gov/statfacts/html/breast.html.

39. Murphy, C. C., Wallace, K., Sandler, R. S. & Baron, J. A. Racial disparities in incidence of young-onset colorectal cancer and patient survival. Gastroenterology 156, 958–965 (2019).

40. Ng, M. C. Y. Genetics of type 2 diabetes in African Americans. Curr. Diab. Rep. 15, 74 (2015).

41. Rostand, S. G., Kirk, K. A., Rutsky, E. A. & Pate, B. A. Racial differences in the incidence of treatment for end-stage renal disease. N. Engl. J. Med. 306, 1276–1279 (1982).

42. https://jamanetwork.com/journals/jamanetworkopen/fullarticle/2788170.

43. https://jamanetwork.com/journals/jamanetworkopen/fullarticle/2835519.

44. Rodosthenous, R. S. et al. Recontacting biobank participants to collect lifestyle, behavioural and cognitive information via online questionnaires: lessons from a pilot study within FinnGen. BMJ Open 12, e064695 (2022).

45. Gajda, M., Kowalska, M. & Zejda, J. E. Impact of two different recruitment procedures (random vs. Volunteer selection) on the results of seroepidemiological study (SARS-CoV-2). Int. J. Environ. Res. Public Health 18, 9928 (2021).

46. Fry, A. et al. Comparison of sociodemographic and health-related characteristics of UK Biobank participants with those of the general population. Am. J. Epidemiol. 186, 1026– 1034 (2017).

47. Divo, M. J., Martinez, C. H. & Mannino, D. M. Ageing and the epidemiology of multimorbidity. Eur. Respir. J. 44, 1055–1068 (2014).

48. Le Couteur, D. G. & Thillainadesan, J. What is an aging-related disease? An epidemiological perspective. J. Gerontol. A Biol. Sci. Med. Sci. 77, 2168–2174 (2022).

49. Mt, 63% N. H. V. T. Colorectal cancer screening* (%), adults 45 years and older by state, 2020. https://www.cancer.org/content/dam/cancer-org/research/cancer-facts-and-statistics/colorectal-cancer-facts-and-figures/colorectal-cancer-facts-and-figures-2023.pdf.

50. Dai, H. et al. Global, regional, and national prevalence, incidence, mortality, and risk factors for atrial fibrillation, 1990-2017: results from the Global Burden of Disease Study 2017. Eur. Heart J. Qual. Care Clin. Outcomes 7, 574–582 (2021).

51. Xi, Y. & Xu, P. Global colorectal cancer burden in 2020 and projections to 2040. Transl. Oncol. 14, 101174 (2021).

52. Gwira, J. A., Fryar, C. D. & Gu, Q. Prevalence of total, diagnosed, and undiagnosed diabetes in adults: United States, August 2021-August 2023. NCHS Data Brief (2024) doi:10.15620/cdc/165794.

53. Rodgers, J. L. et al. Cardiovascular risks associated with gender and aging. J. Cardiovasc. Dev. Dis. 6, 19 (2019).

54. Shapira, N. Women’s higher health risks in the obesogenic environment: a gender nutrition approach to metabolic dimorphism with predictive, preventive, and personalised medicine. EPMA J. 4, 1 (2013).

55. Xie, K. et al. Global, regional, and national burden of chronic kidney disease, 1990-2021: a systematic analysis for the global burden of disease study 2021. Front. Endocrinol. (Lausanne) 16, 1526482 (2025).

56. Chesnaye, N. C., Carrero, J. J., Hecking, M. & Jager, K. J. Differences in the epidemiology, management and outcomes of kidney disease in men and women. Nat. Rev. Nephrol. 20, 7–20 (2024).

57. Khan, A. et al. Genome-wide polygenic score to predict chronic kidney disease across ancestries. Nat. Med. 28, 1412–1420 (2022).

58. Martin, A. R. et al. Clinical use of current polygenic risk scores may exacerbate health disparities. Nat. Genet. 51, 584–591 (2019).

59. Managing differential performance of polygenic risk scores across groups: Real-world experience of the eMERGE Network. The American Journal of Human Genetics 111, 999–1005 (2024).

60. Baptista, P. V. Principles in genetic risk assessment. Ther. Clin. Risk Manag. 1, 15–20 (2005).

61. Wang, J. Y. et al. Three open questions in polygenic score portability. Nat. Commun. 17, 942 (2026).

62. Hughes Halbert, C., et al. Social determinants of family health history collection. J. Community Genet. 7, 57–64 (2016).

63. Khan, A. et al. Polygenic risk alters the penetrance of monogenic kidney disease. Nature Communications 14, 1–10 (2023).

64. Vassy, J. L. et al. The GenoVA study: Equitable implementation of a pragmatic randomized trial of polygenic-risk scoring in primary care. Am. J. Hum. Genet. 110, 1841–1852 (2023).

65. Hao, L. et al. Development of a clinical polygenic risk score assay and reporting workflow. Nature Medicine 28, 1006–1013 (2022).

66. Ge, T. et al. Validation of a trans-ancestry polygenic risk score for type 2 diabetes in diverse populations. medRxiv (2021) doi:10.1101/2021.09.11.21263413.

67. Conway-Pearson, L. S. et al. Family health history reporting is sensitive to small changes in wording. Genet. Med. 18, 1308–1311 (2016).

68. Wilson, B. J. Systematic review: Family history in risk assessment for common diseases. Ann. Intern. Med. 151, 878 (2009).

69. Kirby, J. C. et al. PheKB: a catalog and workflow for creating electronic phenotype algorithms for transportability. Journal of the American Medical Informatics Association : JAMIA 23, 1046 (2016).

70. Shang, N. et al. Medical records-based chronic kidney disease phenotype for clinical care and “big data” observational and genetic studies. npj Digital Medicine **4**, 1–13 (2021).

71. Voss, R. W. et al. Comparing ascertainment of chronic condition status with problem lists versus encounter diagnoses from electronic health records. Journal of the American Medical Informatics Association : JAMIA 29, 770 (2022).

72. Chamberlain, A. M. et al. Identification of Incident Atrial Fibrillation From Electronic Medical Records. Journal of the American Heart Association (2022) doi:10.1161/JAHA.121.023237.

73. Szczerbinski, L. et al. Algorithms for the identification of prevalent diabetes in the All of Us Research Program validated using polygenic scores. Scientific Reports 14, 1–11 (2024).

74. Type 2 Diabetes Mellitus. https://phekb.org/phenotype/type-2-diabetes-mellitus.

75. McLaren, W. et al. The Ensembl Variant Effect Predictor. Genome Biology 17, 1–14 (2016).

76. Khera, A. V. et al. Whole-genome sequencing to characterize monogenic and polygenic contributions in patients hospitalized with early-onset myocardial infarction. Circulation 139, 1593–1602 (2019).

