## Supplemental Methods and Figures for "Evaluation of the genome-informed risk assessment (GIRA) approach from eMERGE in an independent health system"

Supplementary Methods

We restricted individuals to those whose age was between 18 and 75 years at the time of sample collection to ensure that our analysis closely followed the decisions made in the eMERGE Genome Informed Risk Assessment (GIRA) study. When possible, we implemented validated electronic phenotype algorithms from the Phenotype Knowledgebase (PheKb) as this was created by the eMERGE network to create, validate, and disseminate algorithms across different institutions or healthcare systems. However, atrial fibrillation, coronary heart disease, and prostate cancer did not have validated algorithms in the PheKb, so the Polygenic Risk Methods Development Consortium (PRIMED) or the Chronic Condition Warehouse was used.

**Atrial Fibrillation**

The case selection algorithm followed the Chronic Condition Warehouse (CCW) Chronic Condition Algorithm for atrial fibrillation as there was no validated phenotype algorithm in the PheKb. CCW is a medicare-developed validated method to identify individuals with conditions using diagnostic and procedure codes. Details on the algorithm and the ICD-9 and -10 codes used for defining cases can be found here: [ccw-chronic-condition-algorithms.pdf](https://www2.ccwdata.org/documents/10280/19139421/ccw-chronic-condition-algorithms.pdf).

Cases are defined as individuals who have at least one occurrence of an inpatient diagnostic code (ICD9 or ICD10 code) or at least two occurrences of an outpatient diagnostic code at least 30 days apart. A previous study found 1 inpatient or 2 outpatient codes separated by >30 days but within 1 year had a PPV of 78.7% and sensitivity of 92.5% in their prevalent approach while their incident approach had a PVV of 69.7% and sensitivity of 93.6%. This validated the use of the CCW’s algorithms for defining atrial fibrillation cases. Age was calculated using the date associated with the first occurrence of a diagnostic code for atrial fibrillation.

Controls are defined as individuals who had at least two encounters in the system and no occurrence of the atrial fibrillation diagnostic codes. Age was calculated using the date associated with the last recorded encounter in an individual’s electronic health record.

**Breast Cancer**

The case and control selection algorithm followed the PheKb algorithm for breast cancer. We only considered the female case and control algorithm as the GIRA study restricted their analysis to females for breast cancer. Details on the algorithm along with the diagnostic and history codes can be found here: <https://phekb.org/phenotype/breast-cancer>.

Female Breast Cancer Case definition:

- Female
- Any age
- At least one occurrence of the female breast cancer diagnostic codes or at least two occurrences from distinct calendar days of breast cancer history codes if the patient has no breast cancer diagnostic codes

Age was calculated using the date associated with the first occurrence of a female breast cancer diagnostic code or breast cancer history code if the patient had no breast cancer diagnostic code.

Female Breast Cancer Controls definition:

- Female
- Age ≥ 18 years
- None of the female breast cancer diagnostic codes or the breast cancer history codes

Age was calculated using the date associated with the last recorded encounter in an individual’s electronic health record.

**Chronic Kidney Disease**

The case and control selection algorithm was adapted from PheKb algorithm for chronic kidney disease (CKD). The PheKb algorithm relies on CKD staging as we were not interested in the severity of CKD. Instead, we incorporated the simplified CKD case-control definition from this study, <https://www.nature.com/articles/s41467-023-43878-9#Sec8>, which was based on the PheKb algorithm.

**Cases Definition:**

- **Case Definition 1:** received a renal replacement therapy (dialysis or kidney transplant diagnosis or procedure code). If renal replacement therapy was from a dialysis code, then we ensured the code did not co-occur with acute conditions (acute kidney injury, prenatal kidney injury, sepsis, volume depletion, or shock in). The co-occurrence is defined as that the acute condition happens before or after 31 days of the most recent eGFR.
- **Case Definition 2:** no instance of a kidney transplant, no dialysis, at least one instance of eGFR measure, and the most recent eGFR did not co-occur with acute conditions (acute kidney injury, prenatal kidney injury, sepsis, volume depletion, or shock in). The co-occurrence is defined as that the acute condition happens before or after 31 days of the most recent eGFR. After all these criteria were met, the most recent was eGFR below 60 ml/min/1.73m^2^ and one or more instance of CKD or other kidney disease based on diagnostic or procedure billing codes.

For controls, we made sure that they have no instance of a kidney transplant, no dialysis, at least one instance of eGFR measure, and the most recent eGFR did not co-occur with acute conditions (acute kidney injury, prenatal kidney injury, sepsis, volume depletion, or shock in). The co-occurrence is defined as that the acute condition happens before or after 31 days of the most recent eGFR. After all these criteria were met, controls had eGFR greater than 90 ml/min/1.73m^2^ and no evidence of CKD or other kidney disease based on diagnostic or procedure billing codes.

Details on the algorithm along with the diagnosis and procedure codes and lab values used can be found here: <https://phekb.org/phenotype/chronic-kidney-disease>.

The age for cases was calculated using the date associated with the most recent eGFR below 60 ml/min/1.73m^2^ for chronic kidney disease. For controls, age was calculated using the date associated with the last recorded encounter in an individual’s electronic health record.

**Colorectal Cancer**

The case and control selection algorithm was adapted from PheKb algorithm for colorectal cancer. Details on the algorithm along with the diagnosis and procedure codes and colorectal cancer registry tumor site codes used can be found here: <https://phekb.org/phenotype/colorectal-cancer-crc>

Colorectal Cancer Case definition:

- **Case Definition 1:** Evidence from cancer registry that they have been diagnosed with cancer of colon by tumor site code
- **Case Definition 2:** Individuals who failed to qualify as cases by the cancer registry were defined as being a colorectal cancer cases if
  - Had at least one colorectal cancer diagnosis code
  - During the period spanning 365 days before through 365 days after the date of a qualifying colorectal cancer diagnosis code, has at least one procedure code indicating a surgical procedure to treat colorectal cancer
- **Case Definition 3:** Individuals who failed to qualify as cases by definition 1 or 2 then they were defined as being a colorectal cancer cases if
  - Had at least one colorectal cancer diagnosis code
  - During the period spanning 365 days before through 365 days after the date of a qualifying CC diagnosis code, has at least one procedure code for chemotherapy or radiation therapy used to treat CC
  - Has no evidence of any other cancer diagnosis ever
  - Has no evidence of a diagnosis of thrombocytopenia

Age was calculated using the date associated with the first occurrence of a diagnostic code for colorectal cancer.

Controls are defined as individuals who had no CC diagnosis code and had at least one procedure code for colonoscopy. Age was calculated using the date associated with the last recorded encounter in an individual’s electronic health record.

**Coronary Heart Disease**

The case and control selection algorithm followed the coronary heart disease algorithm developed by the PRIMED Consortium. Cases are defined as individuals who have at least one occurrence of the revascularization procedure code or at least three occurrences from distinct calendar days of coronary heart disease hard diagnostic codes if the individual had no history of a revascularization code. Age was calculated using the date associated with the first occurrence of a revascularization procedure code or hard diagnostic code.

Controls are defined as individuals who have at least two encounters in the system and no revascularization procedure code and no hard diagnostic codes. Age was calculated using the date associated with the last recorded encounter in an individual’s electronic health record.

**Hypercholesterolemia**

The case and control selection algorithm followed the hypercholesterolemia algorithm in PheKb. Details on the algorithm along with the diagnosis and procedure codes and measurement and laboratory values used can be found here: <https://phekb.org/phenotype/electronic-health-record-based-phenotyping-algorithm-familial-hypercholesterolemia>.

Cases are defined as individuals >= 18 years old with a lipid profile and meeting the following criteria:

- At least one instance LDL-C and TG measurements, and
- No more than two measurement of TG >= 500 mg/dL, and
- No secondary causes of hypercholesterolemia within 1 year prior to the date of the highest LDL-C level (index date), and
- Had a lipid-lowering treatment within a year to 6 weeks prior to the index date then we calculated a pre-treatment LDL-C level by dividing index LDL-C by 0.7, and
- LDL-C >= 155 mg/dL

Controls were defined as individuals following all the above criteria except:

- LDL-C =< 130 mg/dL
- Individuals whose LDL-C was between 131 and 154 mg/dL were considered as undefined and not included in the analysis.

For controls, age was calculated using the date associated with the last recorded encounter in an individual’s electronic health record. The age for cases was calculated using the date associated with the highest LDL-C level i.e. index date.

**Prostate Cancer**

The case and control selection algorithm was developed by the PRIMED Consortium. We only considered the males for the case and control algorithm as the GIRA study restricted their analysis to males for prostate cancer.

Male Prostate Cancer Case definition:

- Male
- Any age
- At least two occurrences of malignant neoplasm of prostate codes on separate instances

Age was calculated using the date associated with the first occurrence of a malignant neoplasm of prostate code

Male Prostate Cancer Control definition:

- Male
- Age ≥18 years
- At least one encounter
- No occurrences of malignant neoplasm of prostate codes
- No occurrences of non-malignant prostate neoplasm codes
- No prostatectomy codes
- No occurrences of PSA testing

Age was calculated using the date associated with the last recorded encounter in an individual’s electronic health record.

**Type 2 Diabetes**

The case and control selection algorithm followed the Type 2 Diabetes (T2D) algorithm developed by Szczerbinski et al. which modified the Northwestern University algorithm in the PheKb. Details on the algorithm and diagnostic, medication, and laboratory values: <https://www.nature.com/articles/s41598-024-74730-9>.

Type 2 Diabetes cases identification relies on first filtering out individuals with an ICD-based diagnosis of type 1 diabetes. Then, cases were defined using multiple decision paths after ensuring individuals had one or more T2D diagnosis codes:

- **Case Definition 1**: Patient had an outpatient insulin treatment record with no other diabetes medication and at least two different independent ICD diagnosis of type 2 diabetes
- **Case Definition 2**: Patient had an outpatient insulin treatment record with another non-insulin diabetes medication and insulin was prescribed after the non-insulin diabetes medication and at least two different independent ICD diagnosis of type 2 diabetes (i.e. If a patient has an outpatient insulin treatment record, then had prescription of at least one additional non-insulin diabetes medication before initiation of insulin therapy)
- **Case Definition 3**: Patient had no outpatient insulin treatment record but had another non-insulin diabetes medication
- **Case Definition 4:** Patient had no outpatient insulin treatment record and no non-insulin diabetes medication but had abnormal glycemic laboratory results (fasting glucose >= 126 mg/dl or HbA1c >= 6.5%)
- **Case Definition 5:** Patients had no T2D diagnosis codes but had one or more prescriptions for non-insulin diabetes medication and had abnormal glycemic laboratory results (fasting glucose >= 126 mg/dL or HbA1c >= 6.5%)

Age was calculated using the date associated with the first occurrence of a type 2 diabetes diagnostic code, a type 2 diabetes medication, or an abnormal laboratory value

Controls are defined as individuals who have at least one encounters in the system, at least one glucose measurement, no abnormal glycemic laboratory values, no type 2 diabetes diagnostic codes, no instance of any diabetes medication. Age was calculated using the date associated with the last recorded encounter in an individual’s electronic health record.

**Obesity**

The case and control selection algorithm was adapted from the Geisinger Extreme Obesity Algorithm. The algorithm defined cases using various obesity cutoffs, but we instead focused on individuals whose body mass index (BMI) values were above 30 kg/m^2^. Details on the algorithm can be found here: <https://www.ncbi.nlm.nih.gov/projects/gap/cgi-bin/GetPdf.cgi?id=phd004984.1>.

Prior to defining obesity cases, we implemented the following exclusion criteria on BMI measurements:

- Exclude subjects with malignant cancer, which was identified by meeting one of the criteria listed in the algorithm
- Exclude BMI values <14kg/m^2^ or >90 kg/m^2^
- Identify the most recent BMI measure
- Limit the BMI values that occur within the 4 years prior to the most recent BMI measure
- Exclude subjects that had pregnancy code within the 4 years prior to the most recent valid BMI measure
- Calculate the median BMI
- Remove all BMI measure that are >5 kg/m^2^ away fro median BMI
- Remove BMI measures that occur within 30 days of the previous BMI measure
- Exclude subjects that have <3 BMI measures remaining

Obesity Case definition:

- Case Type #1: 100% of BMI measures were >= 30 kg/m^2^
- Case Type #2: Definitely not Case Type #1 and >= 75% of BMI measures >= 30 kg/m^2^

Obesity Control definition:

- Control Type #1: 100% of BMI measures are >=20 kg/m^2^ and <30 kg/m^2^
- Control Type #2: Not Control Type 1 and >=75% of BMI measures are >=20 kg/m^2^ and <30 kg/m^2^

Age was calculated using the date associated with the most recent valid BMI measure that fit the criteria for case and control definition.

Supplementary Figures

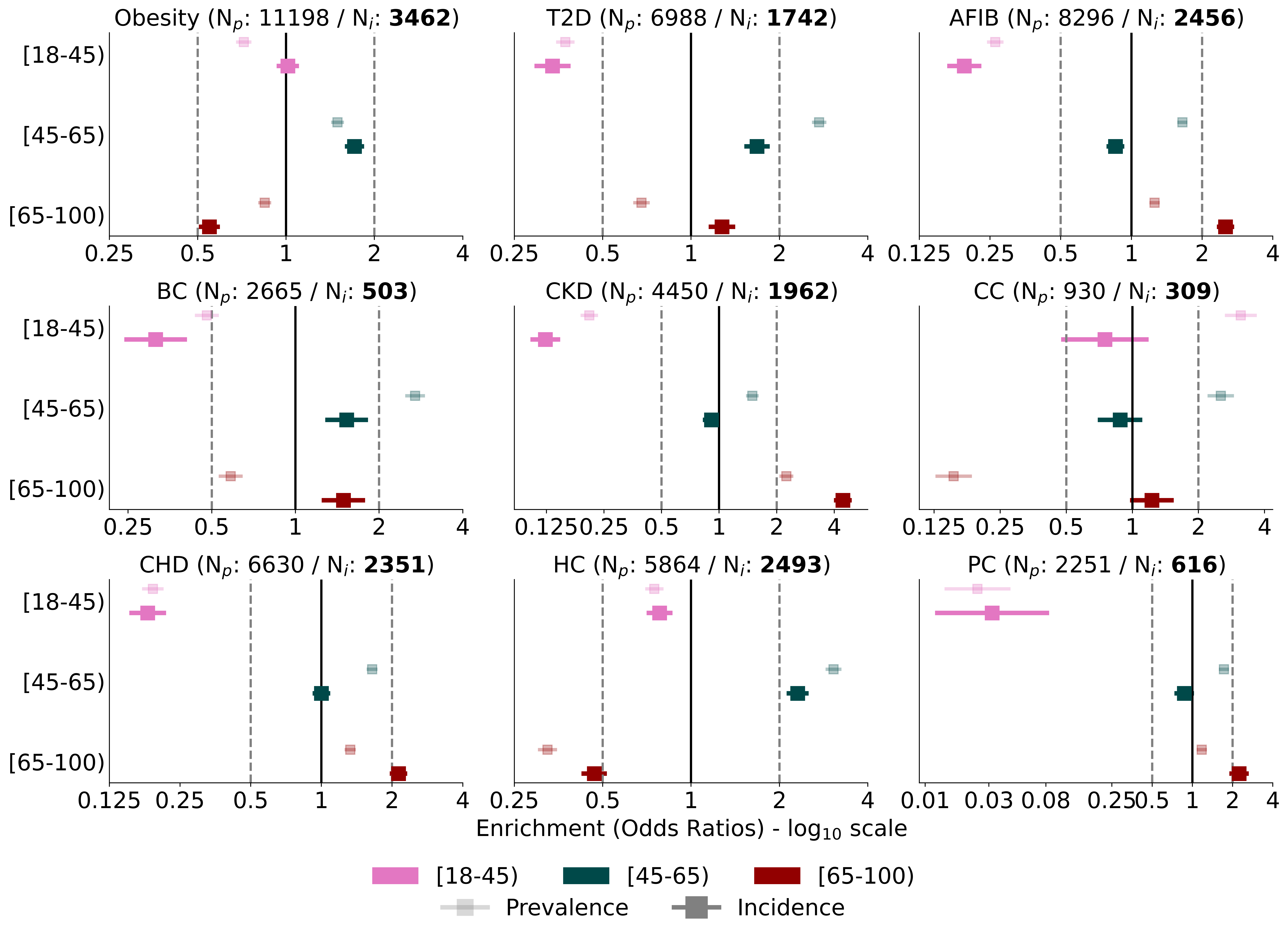

**Supplementary Figure 1.** **Age enrichment within nine conditions as compared to PMBB participants.** Enrichment within prevalent (lighter shade) versus incident (darker shade) cases across the nine adult GIRA conditions is estimated using odds ratios and their standard errors. N_p_ (N_i_) denotes the number of prevalent (incident) cases for each condition among biobank participants aged 18 to 75 at the time of sample collection who were not enrolled in the Penn Medicine Cancer Risk Evaluation Program (N_total_=48,279). Odds ratios are plotted on the log_10_ scale.
Abbreviations: GIRA: Genome Informed Risk Assessment, PMBB: Penn Medicine Biobank, T2D: Type 2 Diabetes, AFIB: Atrial Fibrillation, BC: Breast Cancer, CKD: Chronic Kidney Disease, CC: Colorectal Cancer, CHD: Coronary Heart Disease, HC: Hypercholesterolemia, PC: Prostate Cancer

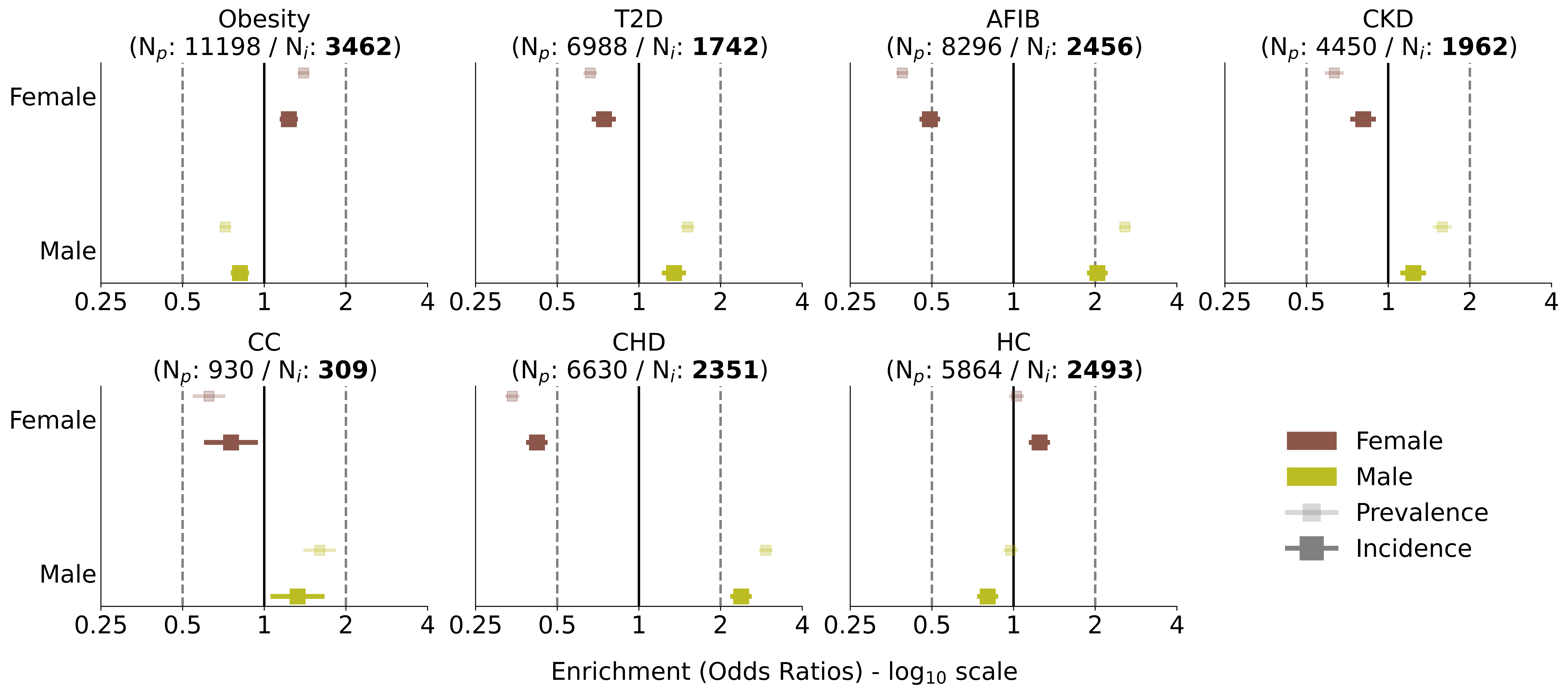

**Supplementary Figure 2.** **Sex enrichment within nine conditions as compared to PMBB participants.** Enrichment within prevalent (lighter shade) versus incident (darker shade) cases across the nine adult GIRA conditions is estimated using odds ratios and their standard errors. N_p_ (N_i_) denotes the number of prevalent (incident) cases for each condition among biobank participants aged 18 to 75 at the time of sample collection who were not enrolled in the Penn Medicine Cancer Risk Evaluation Program (N_total_=48,279). Odds ratios are plotted on the log_10_ scale. Sex enrichments were not plotted for breast cancer and prostate cancer as analyses were restricted to only females and males, respectively.
Abbreviations: GIRA: Genome Informed Risk Assessment, PMBB: Penn Medicine Biobank, T2D: Type 2 Diabetes, AFIB: Atrial Fibrillation, BC: Breast Cancer, CKD: Chronic Kidney Disease, CC: Colorectal Cancer, CHD: Coronary Heart Disease, HC: Hypercholesterolemia, PC: Prostate Cancer

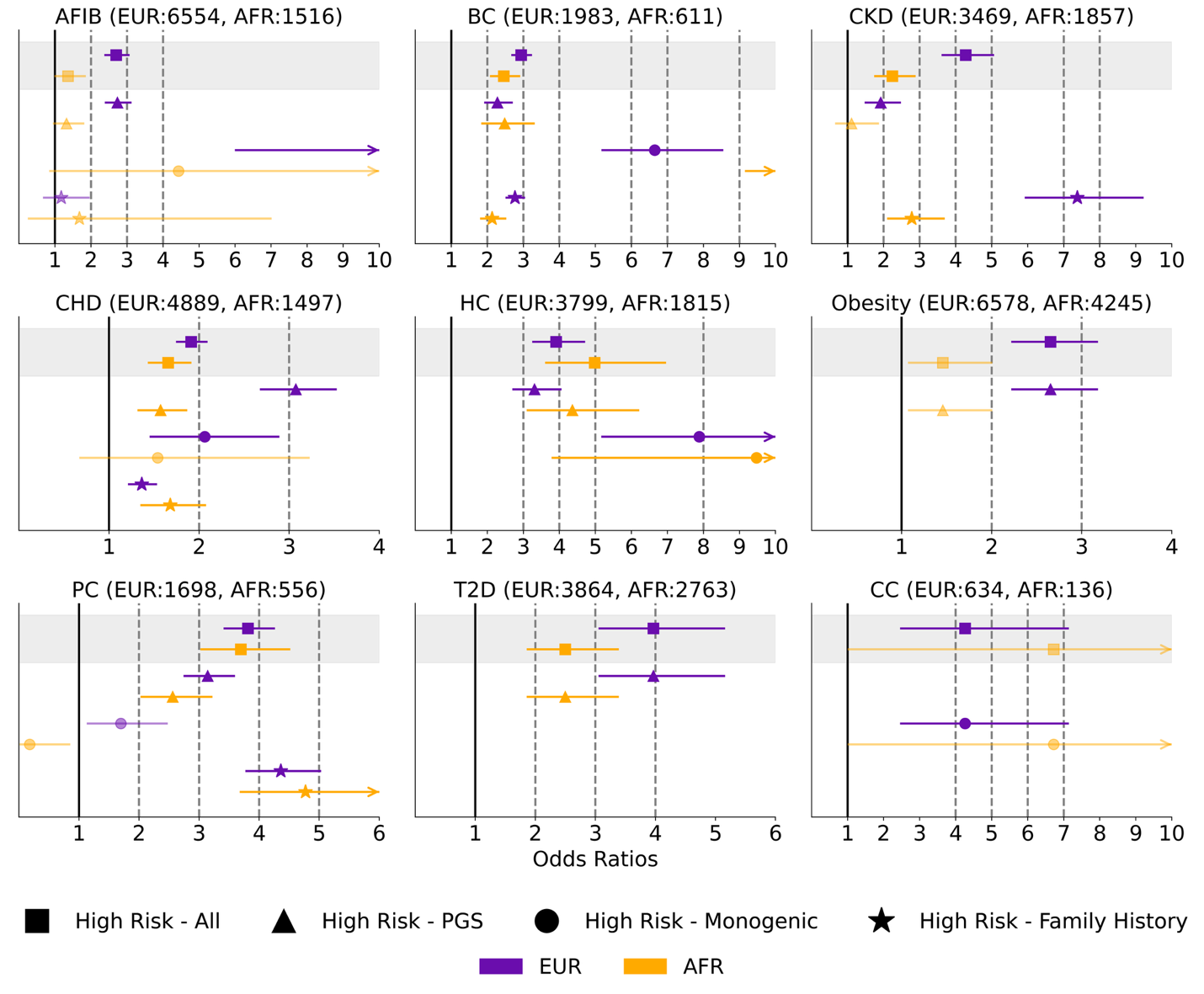

**Supplementary Figure 3. Odds Ratios** **for GIRA high risk stratified by the three genetic risk components of the GIRA report: Polygenic Risk Score (PGS), Monogenic, and Family History.** PMBB participants were labeled as high-risk for each condition using the eMERGE Genome Informed Risk Assessment (GIRA) together with their condition-specific criteria for each risk indicator. All risk indicators are aggregated into a combined risk score to distinguish individuals at high genetic risk, based on the GIRA-recommended criteria for PGS, monogenic risk, and family history, from those without high genetic risk status. High Risk - All models were adjusted for sex, age, age^2^, and first ten genotype or exome PCs. High Risk - PGS models were adjusted for sex, age, age^2^, and first ten genotype PCs. High Risk - Monogenic models were adjusted for sex, age, age^2^, and first exome PCs. High Risk - Family History models were adjusted for sex, age, and age^2^.. 
Abbreviations: T2D: Type 2 Diabetes, AFIB: Atrial Fibrillation, BC: Breast Cancer, CKD: Chronic Kidney Disease, CC: Colorectal Cancer, CHD: Coronary Heart Disease, HC: Hypercholesterolemia, PC: Prostate Cancer

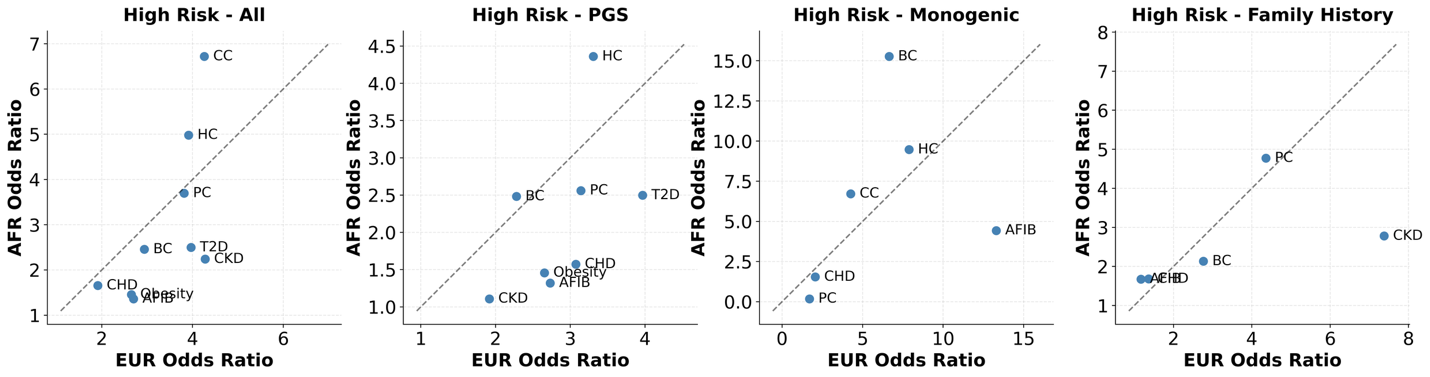

A

B

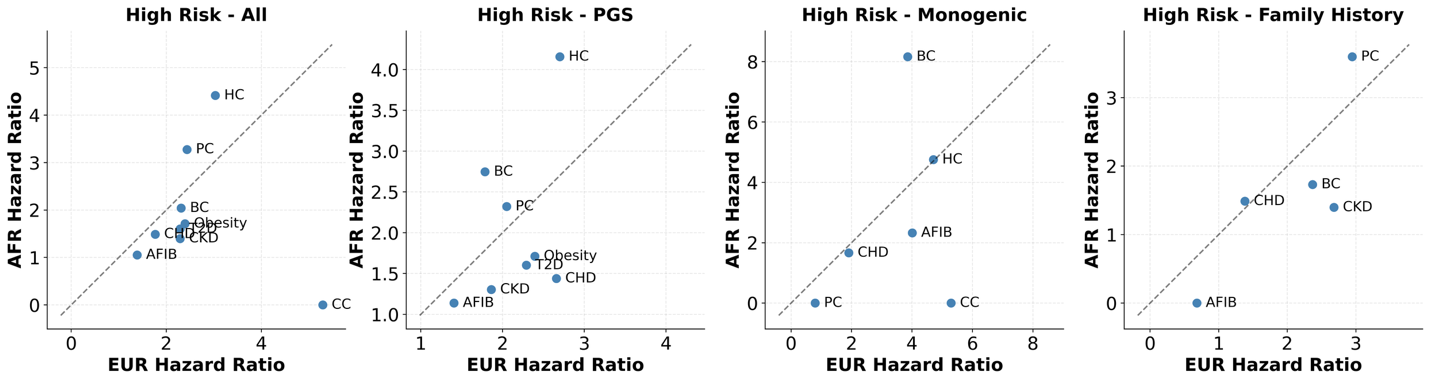

**Supplementary Figure 4. Comparison of odds ratios between EUR and AFR ancestry individuals across genetic risk indicators.** (A) We compare odds ratios between AFR and EUR ancestry individuals for nine adult conditions across each genetic risk component of the GIRA report. (B) We compare hazard ratios between AFR and EUR ancestry individuals for nine adult conditions across each genetic risk component of the GIRA report. Each dot denotes a condition. The diagonal line represents perfect concordance (${OR}_{AFR}= {OR}_{EUR} / {HR}_{AFR}= {HR}_{EUR})$. Points above the line indicate larger effects in AFR populations, while points below indicate larger effects in EUR populations.

Abbreviations: EUR: European American ancestry, AFR: African/African American ancestry, GIRA: Genome Informed Risk Assessment, T2D: Type 2 Diabetes, AFIB: Atrial Fibrillation, BC: Breast Cancer, CKD: Chronic Kidney Disease, CC: Colorectal Cancer, CHD: Coronary Heart Disease, HC: Hypercholesterolemia, PC: Prostate Cancer

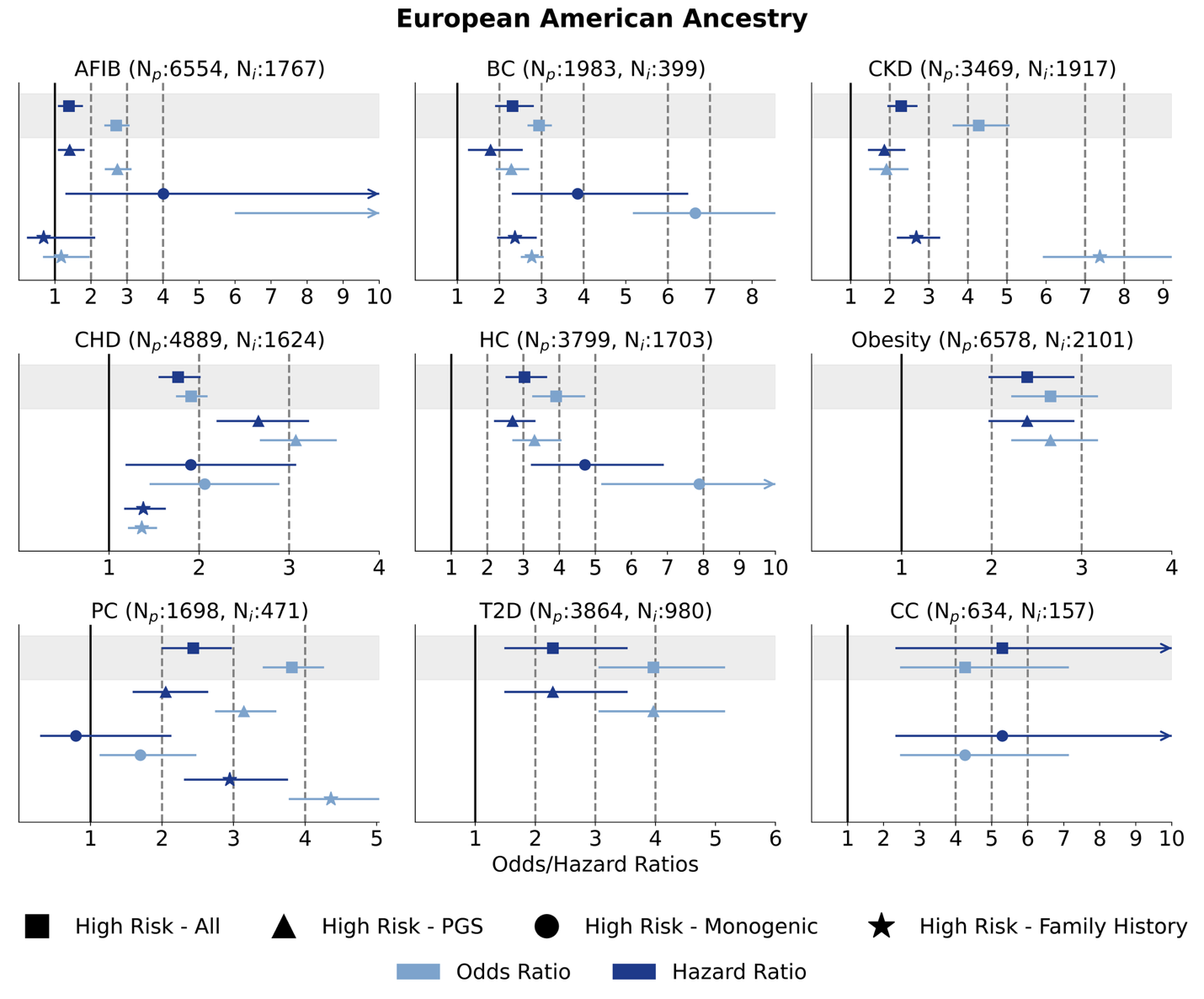

**Supplementary Figure 5.** **Comparison of Odds Ratios and Hazard ratios in EUR ancestry individuals stratified by the three genetic risk components.** Hazard ratios for incident cases were compared to prevalent cases for European American ancestry PMBB participants. Odds/ Hazard ratios for GIRA high risk stratified by the three genetic risk components of the GIRA report: Polygenic Risk Score (PGS), Monogenic, and Family History. High Risk - PGS models were adjusted for sex, age, age^2^, and first ten genotype PCs. High Risk - Monogenic models were adjusted for sex, age, age^2^, and first exome PCs. High Risk - Family History models were adjusted for sex, age, and age^2^. For each hazard ratio model, observation time was defined as the earliest of the following times: date of first diagnosis, last clinical encounter, or death. The disease status was 1 if the individual developed the condition and 0 otherwise.

Abbreviations: T2D: Type 2 Diabetes, AFIB: Atrial Fibrillation, BC: Breast Cancer, CKD: Chronic Kidney Disease, CC: Colorectal Cancer, CHD: Coronary Heart Disease, HC: Hypercholesterolemia, PC: Prostate Cancer

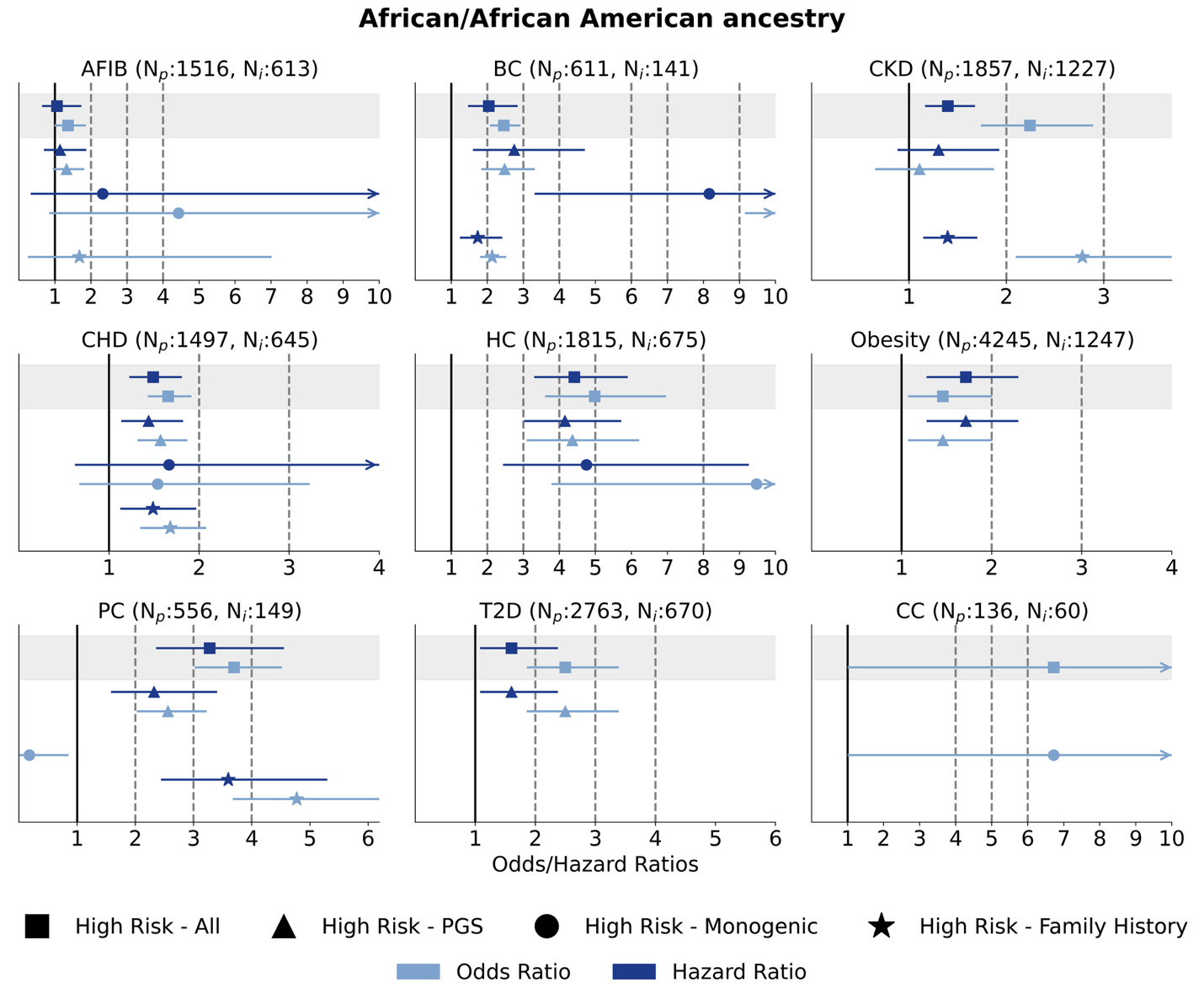

**Supplementary Figure 6.** **Comparison of Odds Ratios and Hazard ratios in AFR ancestry individuals stratified by the three genetic risk components.** Hazard ratios for incident cases were compared to prevalent cases for African/African American ancestry PMBB participants. Odds/ Hazard ratios for GIRA high risk stratified by the three genetic risk components of the GIRA report: Polygenic Risk Score (PGS), Monogenic, and Family History. High Risk - PGS models were adjusted for sex, age, age^2^, and first ten genotype PCs. High Risk - Monogenic models were adjusted for sex, age, age^2^, and first exome PCs. High Risk - Family History models were adjusted for sex, age, and age^2^. For each hazard ratio model, observation time was defined as the earliest of the following times: date of first diagnosis, last clinical encounter, or death. The disease status was 1 if the individual developed the condition and 0 otherwise.

Abbreviations: T2D: Type 2 Diabetes, AFIB: Atrial Fibrillation, BC: Breast Cancer, CKD: Chronic Kidney Disease, CC: Colorectal Cancer, CHD: Coronary Heart Disease, HC: Hypercholesterolemia, PC: Prostate Cancer

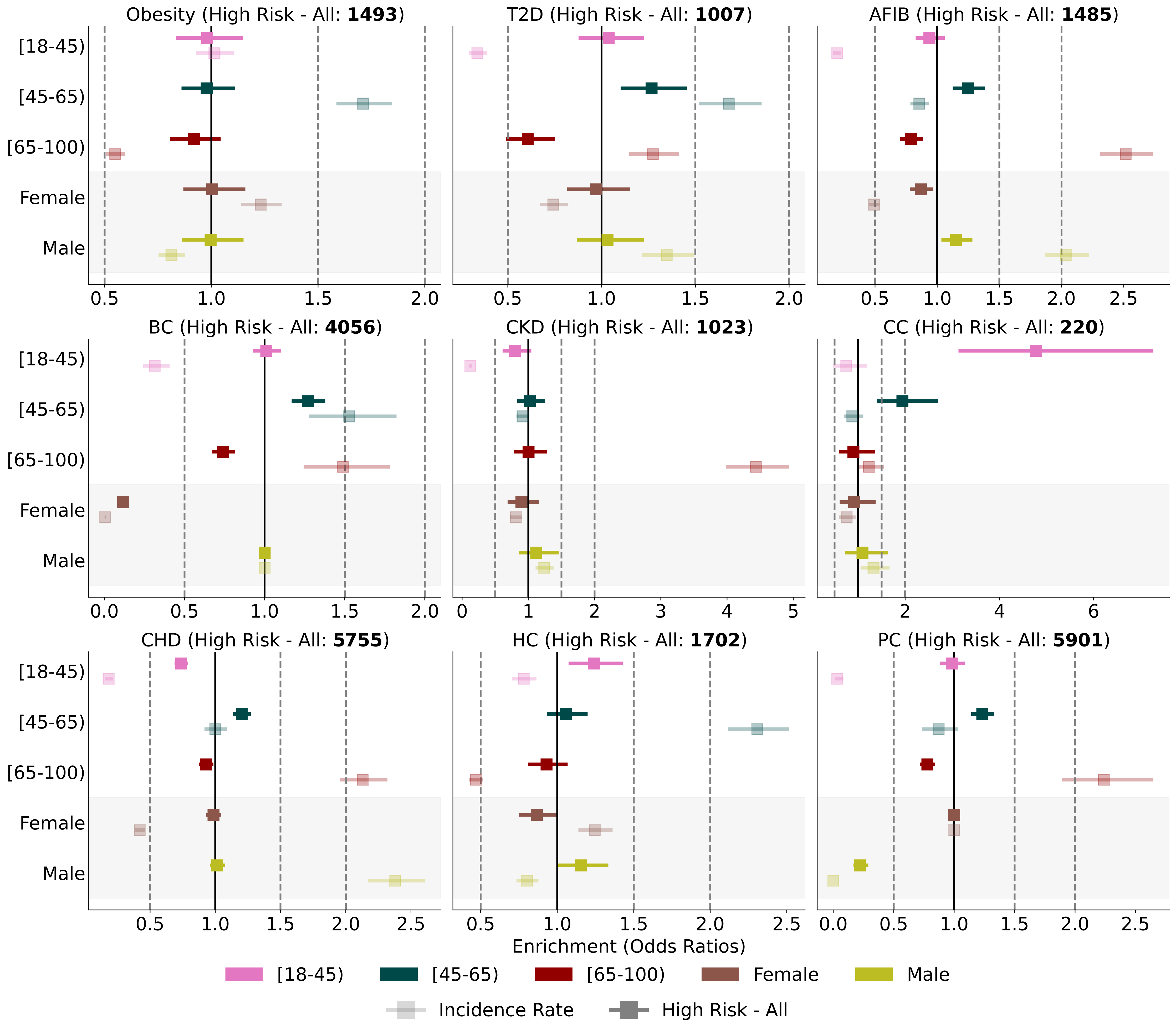

**Supplementary Figure 7. Age and sex enrichments among high-risk PMBB participants across nine conditions compared with overall PMBB enrichments.**
Enrichment within incident (lighter shade) cases versus PMBB participants labeled as high-risk (darker shade) across the nine adult GIRA conditions is estimated using odds ratios and their standard errors. Odds ratios are plotted on the log_10_ scale. Sex enrichments were not plotted for breast cancer and prostate cancer as analyses were restricted to only females and males, respectively.
Abbreviations: PMBB: Penn Medicine Biobank, GIRA: Genome Informed Risk Assessment, T2D: Type 2 Diabetes, AFIB: Atrial Fibrillation, BC: Breast Cancer, CKD: Chronic Kidney Disease, CC: Colorectal Cancer, CHD: Coronary Heart Disease, HC: Hypercholesterolemia, PC: Prostate Cancer

Secondary Analysis Supplementary Figures

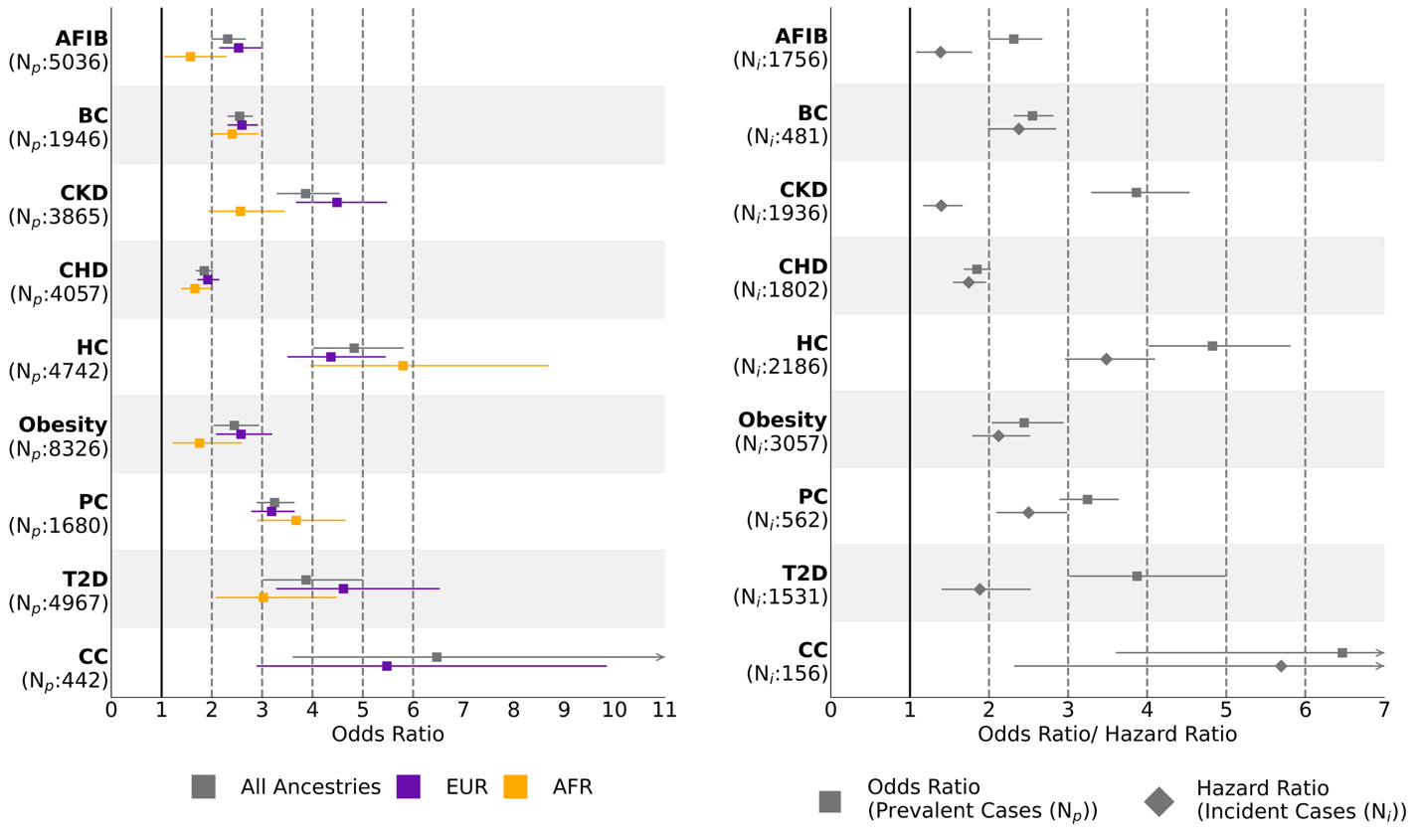

**Supplementary Figure 8. GIRA evaluation in PMBB across nine conditions considered by the eMERGE study for individuals not lost during follow-up.** PMBB participants were labeled as high-risk for each condition using the eMERGE Genome Informed Risk Assessment (GIRA) together with their condition-specific criteria for each risk indicator. All risk indicators are aggregated into a combined risk score to distinguish individuals at high genetic risk—based on the GIRA-recommended criteria for PGS, monogenic risk, and family history—from those without high genetic risk status. Odds/ Hazard ratios were estimated in a model adjusting for sex, age, age^2^, and first ten genotype or exome PCs. For each hazard ratio model, observation time was defined as the earliest of the following times: date of first diagnosis, last clinical encounter, or death. Odds ratios were stratified by ancestry for prevalent cases (left). Hazard ratios for incident cases were compared to prevalent cases for all PMBB participants, irrespective of ancestry (right). We restricted to individuals not lost due during follow-up defined as those who had at least one encounter in the two years prior to the last encounter date.

Abbreviations: PMBB: Penn Medicine Biobank, T2D: Type 2 Diabetes, AFIB: Atrial Fibrillation, BC: Breast Cancer, CKD: Chronic Kidney Disease, CC: Colorectal Cancer, CHD: Coronary Heart Disease, HC: Hypercholesterolemia, PC: Prostate Cancer, EUR: European American ancestry, AFR: African/African American ancestry, AMR: Admixed American ancestry, EAS: East Asian ancestry, SAS: South Asian ancestry

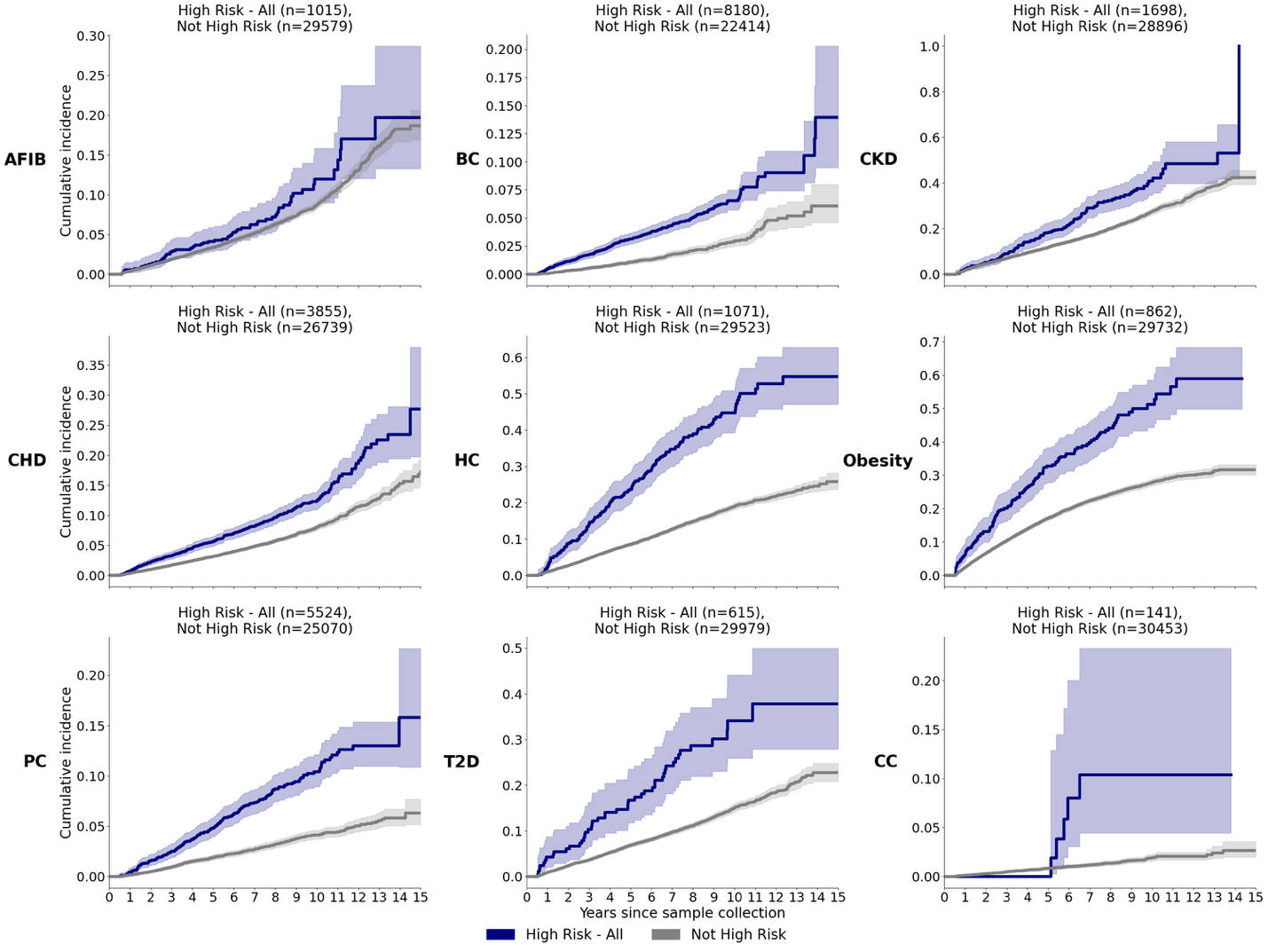

**Supplementary Figure 9. Cumulative incidence curves with stratification by GIRA high-risk versus the rest (‘Not High Risk’) for individuals not lost during follow-up.** Observation time was defined as the earliest of the following times: date of first diagnosis, last follow-up date within a 15-year follow-up window from sample collection. Participants whose high-risk status for each condition was triggered by one or more of the three GIRA genetic risk factors were aggregated into a ‘High Risk - All’ group. We restricted to individuals not lost due during follow-up defined as those who had at least one encounter in the two years prior to the last encounter date.

Abbreviations: GIRA: Genome Informed Risk Assessment, T2D: Type 2 Diabetes, AFIB: Atrial Fibrillation, BC: Breast Cancer, CKD: Chronic Kidney Disease, CC: Colorectal Cancer, CHD: Coronary Heart Disease, HC: Hypercholesterolemia, PC: Prostate Cancer

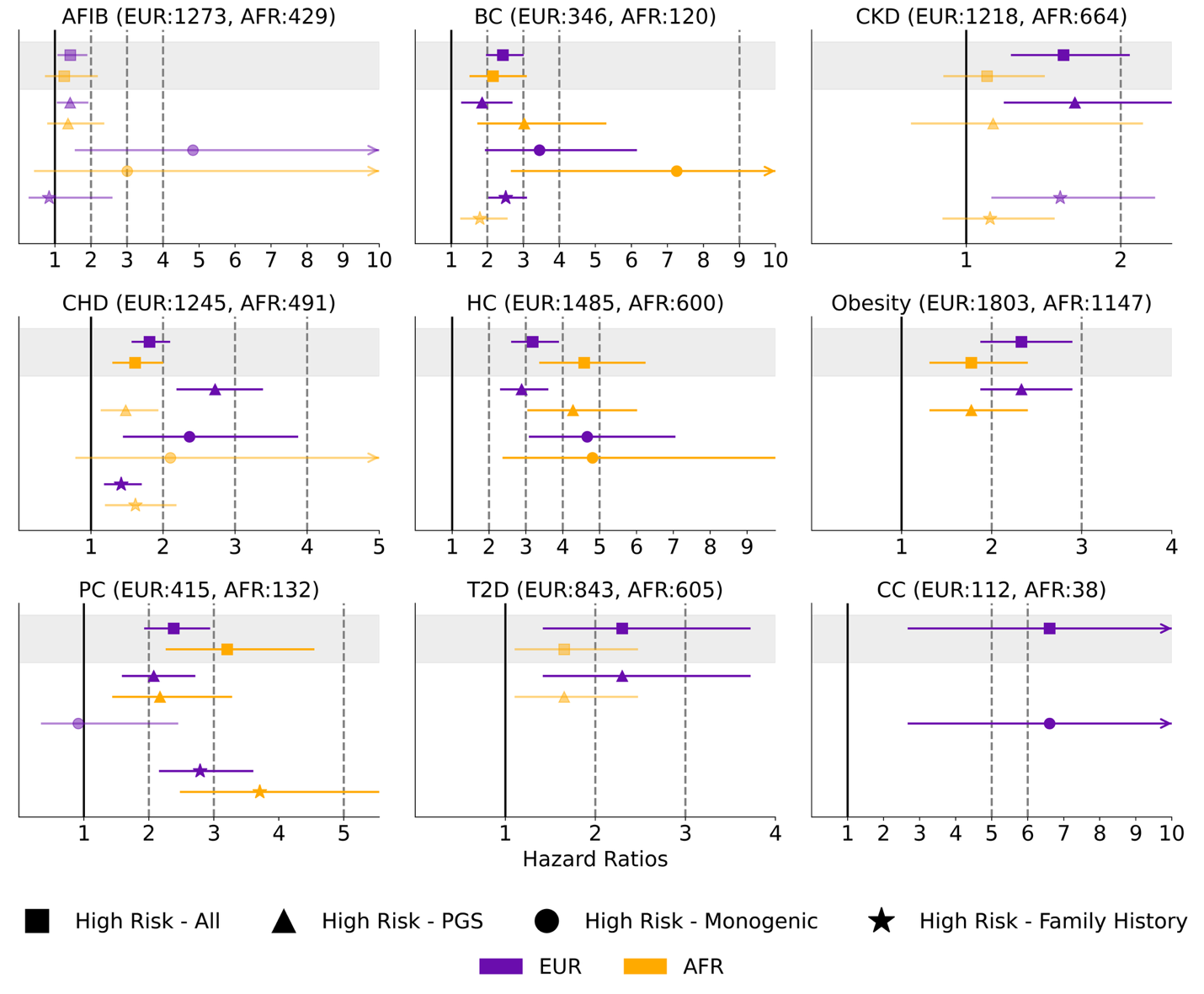

**Supplementary Figure 10. Hazard Ratios** **for GIRA high risk stratified by the three genetic risk components of the GIRA report: Polygenic Risk Score (PGS), Monogenic, and Family History for individuals not lost during follow-up.** High Risk - PGS models were adjusted for sex, age, age^2^, and first ten genotype PCs. High Risk - Monogenic models were adjusted for sex, age, age^2^, and first exome PCs. High Risk - Family History models were adjusted for sex, age, and age^2^. For each model, observation time was defined as the earliest of the following times: date of first diagnosis, last clinical encounter, or death. The disease status was 1 if the individual developed the condition and 0 otherwise. We restricted to individuals not lost due during follow-up defined as those who had at least one encounter in the two years prior to the last encounter date.

Abbreviations: T2D: Type 2 Diabetes, AFIB: Atrial Fibrillation, BC: Breast Cancer, CKD: Chronic Kidney Disease, CC: Colorectal Cancer, CHD: Coronary Heart Disease, HC: Hypercholesterolemia, PC: Prostate Cancer

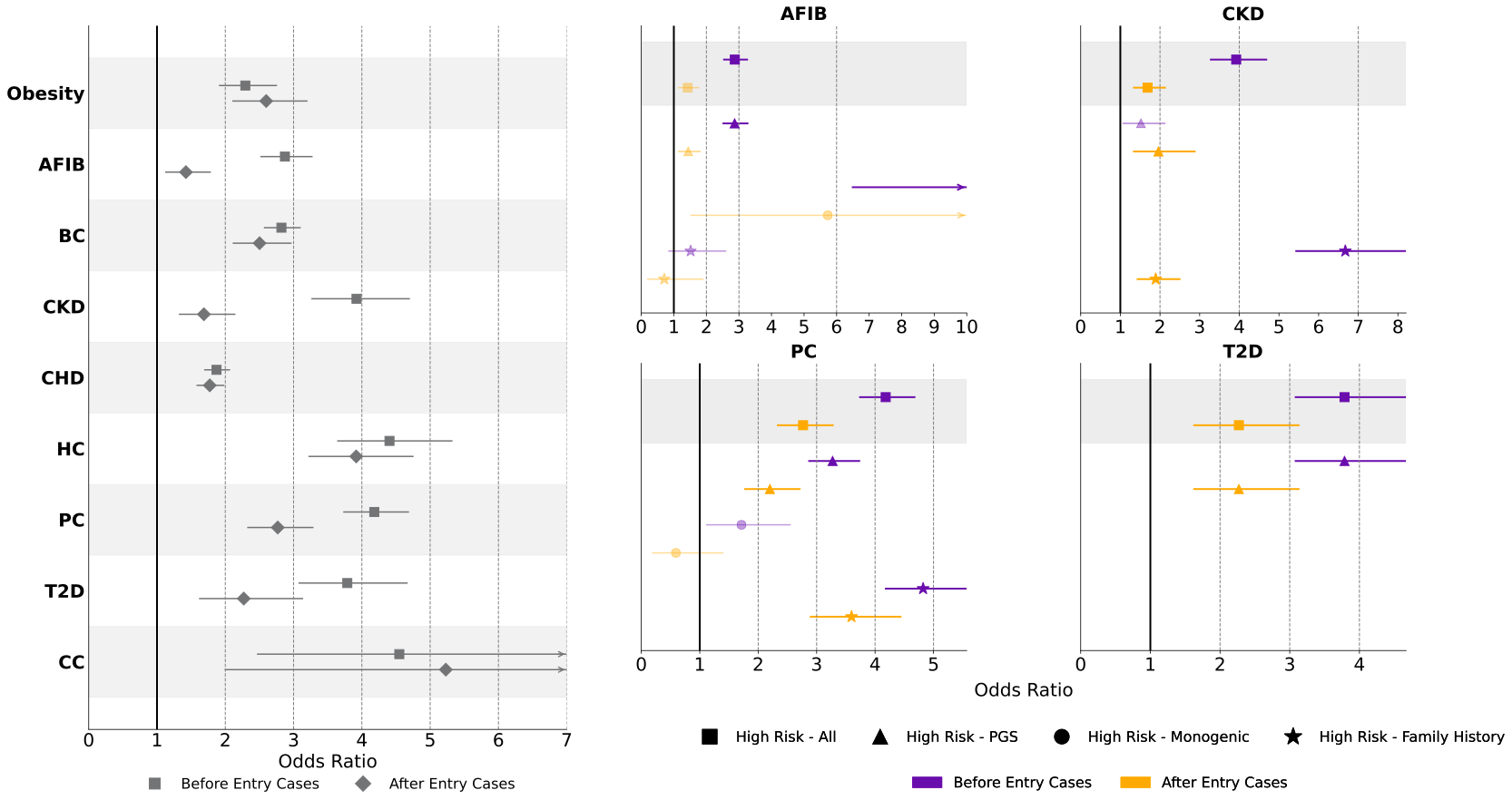

**Supplementary Figure 11. Odds Ratios** **for GIRA high risk stratified by the three genetic risk components of the GIRA report: Polygenic Risk Score (PGS), Monogenic, and Family History for individuals who developed condition before entry compared to individual who developed condition after entry.**

High Risk - PGS models were adjusted for sex, age, age^2^, and first ten genotype PCs. High Risk - Monogenic models were adjusted for sex, age, age^2^, and first exome PCs. High Risk - Family History models were adjusted for sex, age, and age^2^. The disease status was 1 if the individual developed (either before or after entry) the condition and 0 otherwise.

Abbreviations: T2D: Type 2 Diabetes, AFIB: Atrial Fibrillation, BC: Breast Cancer, CKD: Chronic Kidney Disease, CC: Colorectal Cancer, CHD: Coronary Heart Disease, HC: Hypercholesterolemia, PC: Prostate Cancer
